# Nuclear regulatory disturbances precede and predict the development of Type-2 diabetes in Asian populations

**DOI:** 10.1101/2025.02.14.25322264

**Authors:** Pritesh R Jain, Hong Kiat Ng, Darwin Tay, Theresia Mina, Dorrain Low, Nilanjana Sadhu, Ishminder K Kooner, Ananya Gupta, Tai Fei Li, Nicolas Bertin, Calvin Woon Loong Chin, Chai Jin Fang, Liuh Ling Goh, Shi Qi Mok, Su Qin Peh, Charumathi Sabanayagam, Vinitaa Jha, Anuradhani Kasturiratne, Prasad Katulanda, Khadija I Khawaja, Weng Khong Lim, Khai Pang Leong, Ching-Yu Cheng, Jian-Min Yuan, Paul Elliott, Elio Riboli, Lee Eng Sing, Jimmy Lee, Joanne Ngeow, Jian Jin Liu, James Best, Jaspal S Kooner, E-Shyong Tai, Patrick Tan, Rob M van Dam, Woon-Puay Koh, Sim Xueling, Marie Loh, John C Chambers

## Abstract

To identify biomarkers and pathways to Type-2 diabetes (T2D), a major global disease, we completed array-based epigenome-wide association in whole blood in 5,709 Asian people. We found 323 Sentinel CpGs (from 314 genetic loci) that predict future T2D. The CpGs reveal coherent, nuclear regulatory disturbances in canonical immune activation pathways, as well as metabolic networks involved in insulin signalling, fatty acid metabolism and lipid transport, which are causally linked to development of T2D. The CpGs have potential clinical utility as biomarkers. An array-based composite Methylation Risk Score (MRS) is predictive for future T2D (RR: 5.2 in Q4 vs Q1; P=7x10^-25^), and is additive to genetic risk. Targeted methylation sequencing revealed multiple additional CpGs predicting T2D, and synthesis of a sequencing-based MRS that is strongly predictive for T2D (RR: 8.3 in Q4 vs Q1; P=1.0x10^-11^). Importantly, MRS varies between Asian ethnic groups, in a way that explains a large fraction of the difference in T2D risk between populations. We thus provide new insights into the nuclear regulatory disturbances that precede development of T2D, and reveal the potential for sequence-based DNA methylation markers to inform risk stratification in diabetes prevention.

## INTRODUCTION

Type 2 diabetes (T2D) is an age-related, chronic disease typically characterised by the presence of adiposity, insulin resistance and impaired glucose metabolism. Although an important public health problem in all regions of the world, the burden of diabetes is particularly high amongst people from Asian countries^1^. Currently, around 55% of people with T2D live in Asia, and the number of people living with T2D in the Asia-Pacific region is predicted to reach 412 million by 2045^2^. In urban and migrant settings, one in five Asian adults is living with diabetes^3–5^, with disease onset occurring up to 10 years earlier and at substantially lower levels of adiposity, compared to people of European background^6,7^. Understanding the mechanisms underlying susceptibility to diabetes, and improving metabolic health outcomes, amongst Asian people is a major biomedical research priority^8^. As a multifactorial disease, T2D arises through the intersection of a complex genetic background, with multiple modifiable risk exposures, including early life adversity, unfavourable lifestyle behaviours and adipose expansion^9^. We previously hypothesised that the convergence of these exposures would manifest through perturbations of nuclear regulation, that disrupt cellular metabolism and foster the development of diabetes^4^. In keeping with this, we and others identified changes in DNA methylation, a key regulator of DNA conformation, chromatin structure and gene expression, at *ABCG1*, *SREBF1*, *TXNIP* and several other genomic loci, that precede and predict development of T2D by more than a decade^4,10–14^. However, modest sample size, low coverage of DNA methylation profiling and the parsimonious nature of the accompanying clinical and molecular profiling, limited the impact of these observations, either as functional molecular insights or as the basis for biomarker discovery.

To extend on this prior work, and advance understanding of the nuclear and molecular disturbances underlying development of diabetes in Asian populations, we now report comprehensive molecular phenotyping of Asian people at risk for future diabetes, including through application of whole genome and targeted methylation sequencing. In so doing, we identify co-ordinated immune and metabolic pathway disturbances that precede and predict the development of diabetes, and reveal the potential for methylation profiling to improve identification of susceptible individuals.

## RESULTS

Our study design is summarised in **Extended Data Figure 1**. We first carried out epigenome-wide association for incident T2D using the baseline samples collected at enrolment for 5,709 Asian individuals, from three prospective population-based cohorts (**Supplementary Online Materials; Supplementary Table 1**). Diabetes was defined as i. physician diagnosis of diabetes and on drug treatment, ii. fasting glucose ≥7.0mmol/L or iii. HbA1c ≥6.5%^1^. All participants were confirmed free from diabetes at enrolment. Cases (N=2,592) were selected as people identified to have developed diabetes during follow-up of up to 15 years. Controls (N=3,117) were people confirmed to remain free from diabetes during follow-up, and were matched to cases based on age, sex, and ethnic group. DNA methylation was quantified in genomic DNA from whole blood, using the Illumina MethylEpic array (N∼850K markers) in the iHealth-T2D and MEC cohorts^15^, or the Illumina HumanMethylation 450K array (N∼450K markers, LOLIPOP cohort). Epigenome wide association was carried out in the three cohorts separately, followed by inverse-variance meta-analysis (see **Methods**). Statistical significance was inferred as P<8.62x10^-8^, based on permutation testing^16^.

### Epigenome-wide association identifies multiple genetic loci predicting T2D

We identified 420 CpG sites that are associated with well characterised incident T2D at P<8.62x10^-8^; a ∼20-fold increase compared to prior knowledge (**Supplementary Table 2**; **Figure 1**). The CpGs are distributed between 314 genetic loci. Conditional analyses at each locus identified 323 CpG sites to be associated with T2D, independent of other nearby CpGs sites (‘Sentinel CpGs’; **Supplementary Table 2)**. For 306 genetic loci, there was a single Sentinel CpG; at the remaining 8 loci there were 2 or more CpGs independently associated with incident T2D. Twenty-one (6%) of the 323 Sentinel CpGs have been previously reported to be associated with incident T2D. Relative risk (RR) for new onset T2D ranged from 1.11 to 2.51 between the highest and lowest risk quartiles of DNA methylation for the respective markers (**Figure 1**). The mean absolute difference in methylation level between incident T2D cases and controls at the 323 Sentinel CpGs was 0.5%, with no evidence for heterogeneity of effect across Asian ethnic group or between cohorts (**Supplementary Table 3**). After adjusting for glucose and HbA1c, 56 loci remained significantly associated with incident T2D at P<8.62x10^-8^ (**Supplementary Table 4, Supplementary** Figure 1). As a sensitivity analysis, we excluded 1,752 individuals with prediabetes at baseline (HbA1c≥6% or fasting glucose≥6mmol/L); despite reduced sample size, 23 sentinel CpGs remained independently associated with future T2D (P<8.62x10^-8^; **Supplementary Table 4, Supplementary** Figure 1). Our results thus identify extensive variation in DNA methylation that precedes and predicts T2D, including multiple methylation markers that are independent of traditional glycaemic measures of metabolic dysregulation.

**Figure 1.**
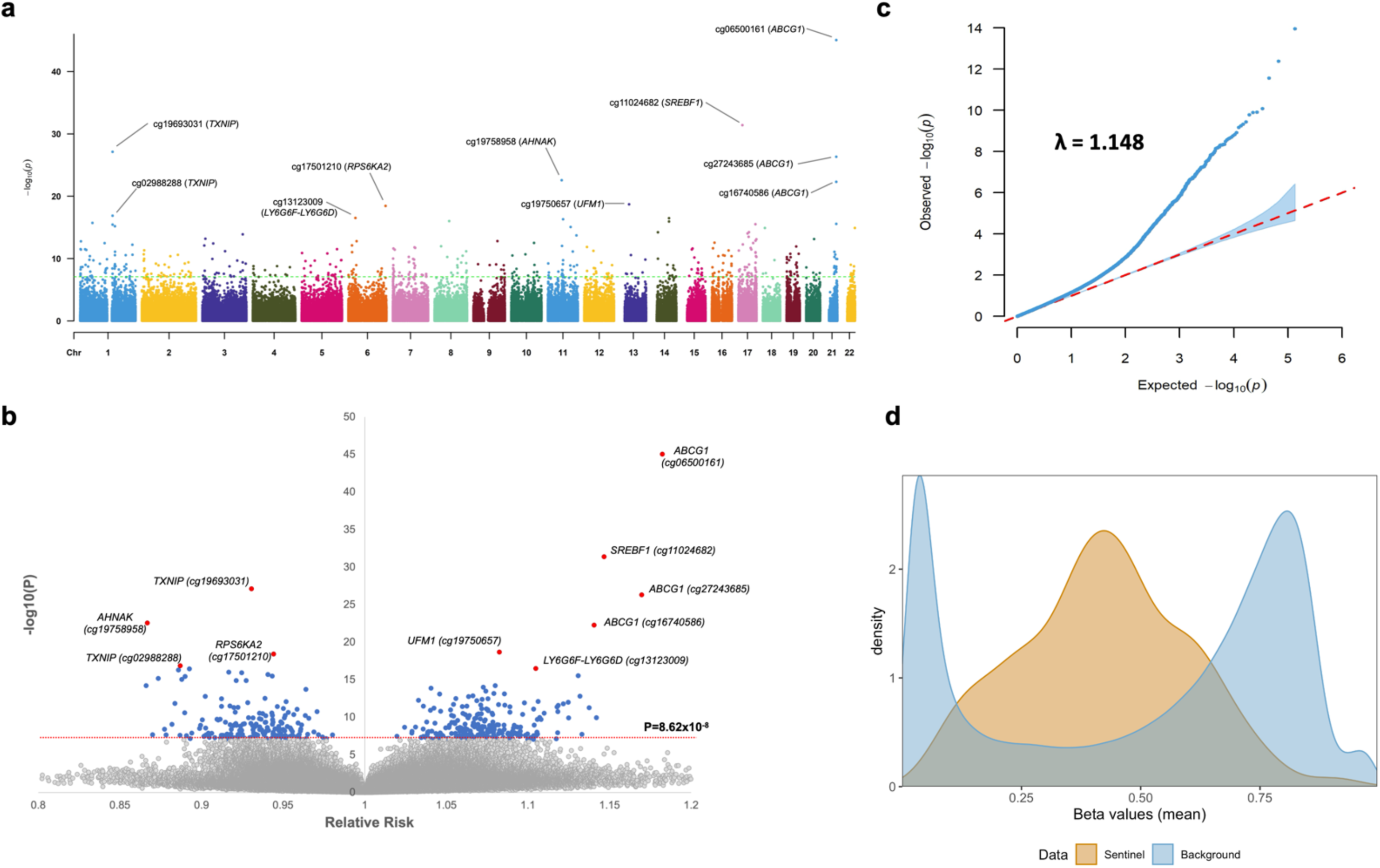
Epigenome Wide Association Study (EWAS) for incident T2D in Asians. **Panel a)** Manhattan plot summarizing the association between DNA methylation at the ∼850K CpG sites assayed in EWAS, and incident T2D. The 10 top-ranking CpGs are annotated with CpG ID and nearest gene. b) Volcano plot showing Relative Risk for incident T2D per 1% change in DNA methylation at the ∼850K CpG sites assayed. **c)** QQ-plot of the p-value for association with incident T2D in the EWAS analysis. Lambda is the genomic control inflation factor. **d)** Density plot showing the distribution of mean methylation levels at the 323 Sentinel CpGs, compared to background CpGs on the array.

### Determinants of DNA methylation at the Sentinel CpG sites

We used data from an independent population cohort comprising 2,237 Asian individuals with deep clinical and molecular phenotyping (Health for Life in Singapore study^17^, HELIOS; **Methods, Supplementary Table 1**), to explore the potential genetic and environmental factors influencing methylation at our Sentinel CpG sites. We tested both directly measured exposures, as well as exposures genetically inferred by polygenic risk scores (PRS), for association with methylation at Sentinel CpGs. In phenome-wide analyses, we observed that the Sentinel CpGs are associated with multiple measures of adiposity and lipid metabolism (range 59-74% of CpGs associated with trait at P<0.05; Fold Enrichment [FE]: 5.4 to 7.1, P_perm_<0.001 compared to background expectations; **Extended Data Figure 2**; **Supplementary Table 5, Supplementary Table 6**). We also found that the Sentinel CpGs are associated with dietary quality, including protective impacts of Alternate Mediterranean, DASH and Healthy Eating Index scores^18–20^, as well as with intakes of fruit, vegetables and other food items as assessed through Asian dietary metabolite panels^21^ (range 10 to 56% of CpGs associated with trait at P<0.05; FE: 1.9 to 6.8, P<0.001 compared to background). Polygenic risk scores for adiposity, lipid metabolism, diabetes and alcohol intake are associated with changes in methylation that are unfavourable for T2D risk, while polygenic risk for educational attainment is protective (**Extended Data Figure 2**; **Supplementary Table 7**). The results of these phenome-wide and polygenic risk analyses support the view that disturbances in methylation at the CpG sites predicting T2D are directly influenced by educational attainment, unfavourable dietary patterns, increased adiposity, lipid disturbances, and genomic disturbances recognised to determine T2D.

### Functional genomic evaluation of the Sentinel CpGs

Multiple lines of evidence point to a role for the identified CpG sites in genome regulation and transcriptional control. First, compared to background CpGs, our Sentinel CpG sites are more likely to show intermediate levels of methylation (79% vs 38%, FE=2.10; P_chi-sq_<5x10^-324^; **Figure 1**), and to have greater variability in methylation (Mean SD: 4.76% vs 4.20%, P_t-test_<5x10^-324^). Second, we show that there is enrichment of the sentinel CpGs compared to background for location in (i) DNase I hotspots (FE: 1.23 to 1.49), (ii) activating histone marks H3K4me1 and H3K36me3 (FE: 1.16 to 4.42), and (iii) Transcription flanking, enhancer, and genic enhancer regions (FE: 1.56 to 4.84) across multiple tissue and cell subsets (FDR – P_Hyper_<0.05, **Supplementary Table 8; Extended Data** Figure 3). Third, we demonstrate that the sentinel CpGs are significantly enriched for location in the binding sites of 92 specific nuclear transcription factors, including ZNF597, NFKB2, SPI1, IKZF1/2, JUNB, IRF4, and CREBBP (FE: 1.21 to 8.07, FDR-P_Hyper_<0.05; **Supplementary Table 9**; **Extended Data Figure 3**). Fourth, we show that the sentinel methylation loci are enriched for association with gene expression in both *cis-* and *trans-* (expression quantitative trait methylation loci [eQTMs], assessed by RNA-seq of whole blood in 1,228 Asian samples; *cis-*eQTMs FE: 4.70, P_perm_<0.001; *trans-*eQTMs FE 1.69, P_perm_=0.03; **Supplementary Table 10**; **Supplementary Table 11**, **Extended Data Figure 3**). Fifth, the sentinel CpGs, and their associated *trans-*gene expression signatures show covariation that is greater than expected under the null hypothesis, consistent with a co-ordinated functional genomic response (**Extended Data Figure 4**). Finally, Gene Set Enrichment Analysis (GSEA) of the *cis-*eQTMs reveals they are enriched for insulin signalling and cholesterol metabolism (in particular through *ABCG1, SREBF1 DBCR24, MSMO1, BAIAP1 and PDK4*; P<0.001; **Supplementary Table 10**, **Supplementary Table 12, Supplementary** Figure 2), while the *trans*-eQTMs are strongly enriched for immune system activation, interferon and cytokine signalling (P<10^-17^; **Supplementary Table 11**, **Supplementary Table 13, Supplementary** Figure 3), coherent biological pathways linked to diabetes and related phenotypic disturbances. Our functional genomic analyses thus strongly support the view that the Sentinel CpGs identify multiple disturbances of nuclear regulation, that precede the development of T2D and related metabolic phenotypes.

### Cis- and trans-mQTLs support causal relationships between DNA methylation, T2D and related metabolic phenotypes

To help understand the causal pathways linking our Sentinel CpGs to diabetes, we next carried out genome-wide-association amongst 7,573 Asian individuals with whole genome sequence and MethylEpic array data (see **Methods**). This enabled us to identify genetic variants that influence our Sentinel CpGs (‘mQTL’ SNPs), which we then used as genetic instruments in Mendelian randomisation, pathway, and related functional genomic analysis. We found *cis*-acting genetic variants influencing 264 (82%) of our Sentinel CpGs (**Supplementary Table 14**), with the sentinel *cis*-mQTLs accounting for a median 1.2% of variation in methylation (range 0 to 24%). We also found 242 genetic loci that influence methylation at 253 (78%) of our Sentinel CpGs in *trans* (**Supplementary Table 15**). Overall, the median total genetic variance explained for the sentinel CpGs was 7.5% (range: 0 to 79%). Both the *cis- and trans-*acting mQTL SNPs are strongly enriched for association with diabetes, adiposity, insulin resistance and lipid concentrations in published GWAS studies^22–31^ (FE: 1.33 to 19.23; P_perm_: 0.05 to <0.001) (**Extended Data Figure 5**, **Supplementary Table 16, Supplementary Table 17**). Furthermore, Summary data-based Mendelian Randomization^32^ (SMR) together with colocalization^33^ (**Methods**), supports the view that our Sentinel CpGs, share a common genetic basis with multiple metabolic, inflammatory and cardiovascular traits and diseases (**Supplementary Table 18** and **Supplementary Table 19**).

Amongst the *cis-*regulated methylation loci, sentinel CpGs at *BAIAP2*, *CPNE6*, *H1-10*, *HMGA1, INAFM2*, *JARID2, MDM4*, *RPS6KA2*, *SLC12A2* and *TGM4* may be causally linked to the development of T2D (SMR P<1x10^-5^ and Coloc Posterior Probability [Coloc PP.H3 or Coloc PP.H4]>0.6; **Extended Data Figure 6; Supplementary Table 18**). We also observed shared genetic regulation between Sentinel CpGs and adiposity (N=8 CpGs), lipid traits (N=33 CpGs), and risk of cardiovascular disease (N=6 CpGs) (**Supplementary Table 18**). This includes an association between methylation at *ABCA1* locus with LDL cholesterol, total cholesterol, and triglycerides (Coloc PP.H3>0.99), and at homeobox *HOXB1* with WHR (Coloc PP.H4>0.97).

Amongst the 242 *trans-*acting mQTLs, SMR also supported the presence for a shared genetic basis between *trans*-methylation and diabetes at (13 *trans-*mQTLs and 45 CpGs, with SMR P<6.6x10^-5^; **Supplementary Table 19**). This includes the *FADS2, NFKB1* and *TBX6* SNP loci, impacting methylation at the *SREBF1*, *CDKAL1*, *DHCR24* and *IGFBP6* loci, and influencing risk of diabetes. The *trans-*acting SNPs also provide evidence for a shared genetic basis between our Sentinel CpGs and adiposity, lipid metabolism, systemic inflammation and cardiovascular risk. This includes the *trans*- effects of SNPs at the *APOA5*, *APOE3*, *FADS*, *NFKB1*, *REST* and *SCARB1* loci (**Supplementary Table 19**). Together, this provides further evidence that the methylation disturbances identified represent components of biological pathways causally linked to the pathophysiology of metabolic disease and its cardiovascular complications.

### The trans-acting mQTLs reveal co-ordinated nuclear regulatory pathways linked to inflammation and metabolic disease

Amongst the 242 *trans-*acting mQTLs, we find that 50 are pleiotropic and influence multiple independent Sentinel CpG sites (range 2 to 45 Sentinel CpGs per SNP locus; FE: 5 to 1729 compared to background, P_Hyper_<2.1x10^-4^; **Figure 2**, **Supplementary Table 20**), indicating an effect on methylation within co-ordinated nuclear pathways. We searched the eQTLgen database^34^ to identify the *cis-*genes and RNA species, that might be molecular mediators for the *trans-*acting effects of these 50 pleiotropic genomic loci. We found 153 (115 protein coding) *cis-*eQTLs that are associated with 33 of our 50 Sentinel SNPs at P<1.2x10^-5^ (i.e. P<0.05 after correction for 4,283 independent tests; **Supplementary Table 21**). Colocalization confirms that 151 (99%) of these *cis*-eQTLs are linked to the respective *trans*-methylation signature (Coloc PP.H3>0.9 or PP.H4>0.9). Our Asian-specific RNA-seq dataset replicated 53 of these colocalising *cis-*eQTLs (P_GWAS_<0.05; N=1,228 samples; 105 eQTLs present in data; **Supplementary Table 21**).

**Figure 2.**
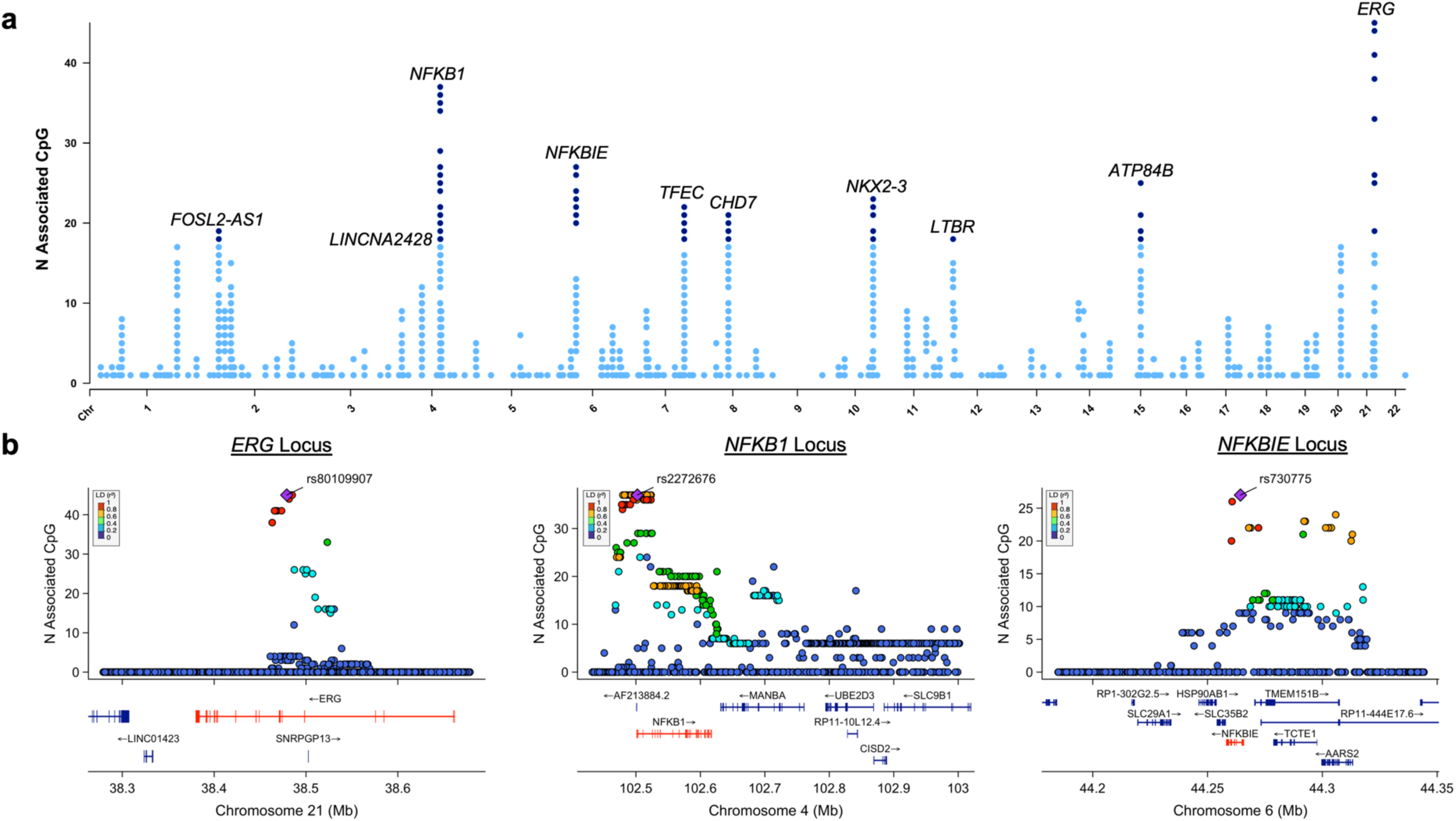
Genome wide analysis of *trans-*acting mQTL SNPs. **Panel a)** The number of Sentinel CpGs (y axis) associated with the genome-wide SNP variation (x axis). Results show that there are discrete genomic regions, characterized by the presence of sequence variation that influences multiple Sentinel CpGs in *trans*. The 10 top-ranking Sentinel SNPs that influence the highest number of Sentinel CpG sites in *trans* (N CpGs≥18) are highlighted in dark blue and annotated by nearest genes to the Sentinel SNP. **b)** Regional plots for the same analysis, showing results at the *ERG*, *NFKB1* and *NFKBIE* loci, the three genomic regions influencing the highest numbers of Sentinel SNPs in *trans*.

We next used hierarchical clustering to organise the *trans-*acting relationships between SNP, gene expression and DNA methylation, into potential biological networks. We focused on the 33 Sentinel *trans*-mQTL SNPs with at least one *cis-*eQTL and identified seven discrete clusters based on their shared relationships to Sentinel CpGs (**Supplementary** Figure 4; see **Methods**). The largest two networks comprise 19 *trans*-acting SNPs linked to 113 colocalising *cis*-eQTLs, 128 *trans*-CpGs and their proximal genes (‘CPG-Genes’; **Extended Data Figure 7; Supplementary** Figure 5). The *cis*-eQTLs mediating these transacting pathways are strongly enriched for transcription factors known to be critical components of inflammatory pathways, and their specific interacting proteins, as well as to key components of metabolic pathways directly relevant to the pathophysiology of diabetes (Count: 1-61 *cis*-eQTLs, FE: 1.26 to 263.9, P_FDR_ > 0.05) (**Extended Data Figure 7**; **Supplementary Table 22)**

To illustrate the complex relationships between immune activation, nuclear regulation and diabetes, we summarise our analyses focused on the *NKFB1* cluster (**Figure 3**). We show that *NFKB1* expression is closely correlated with both measured adiposity (BMI: Beta=0.143 per SD, P=6.2x10^-6^) and genetically inferred adiposity (PRS-BMI: Beta=0.068 per SD, P=0.03), raising the initial epidemiological possibility that *NFKB1* activation might represent a pathway linking excess accumulation of fat to future diabetes (**Figure 3a**, **Figure 3b**). However, we show that sentinel SNP rs2272676 (GàT) which increases *NFKB1* expression in *cis* (eQTLGen Z=15.4), and lowers methylation at 37 Sentinel T2D CpG sites in *trans* (Beta: -0.002 to -0.031 per allele copy, P= 4.1x10^-8^ to 1.8x10^-205^, **Supplementary Table 15**), is associated with a *lower* risk for diabetes (Odds ratio: 0.986 per allele copy, P=7.8x10^-6^, **Supplementary Table 17**). SMR Analysis using rs2272676 confirms a shared genetic basis between increased *NFKB1* expression and lower *trans-*CpG DNA methylation (SMR Beta: -0.35 to -1.99, P=2.4x10^-7^ to 6.1x10^-43^ ), as well between *trans-*CpG DNA methylation and risk of diabetes (SMR Beta: 0.44 to 7.29, P-value: 4.9x10^-4^ to 8.8x10^-6^; **Supplementary Table 19, Supplementary Table 21, Figure 3c**). Our findings suggest that *NFKB1* activation in the context of increased BMI, may in fact ameliorate, and protect from, the adverse metabolic effects characteristic for excess adiposity.

**Figure 3.**
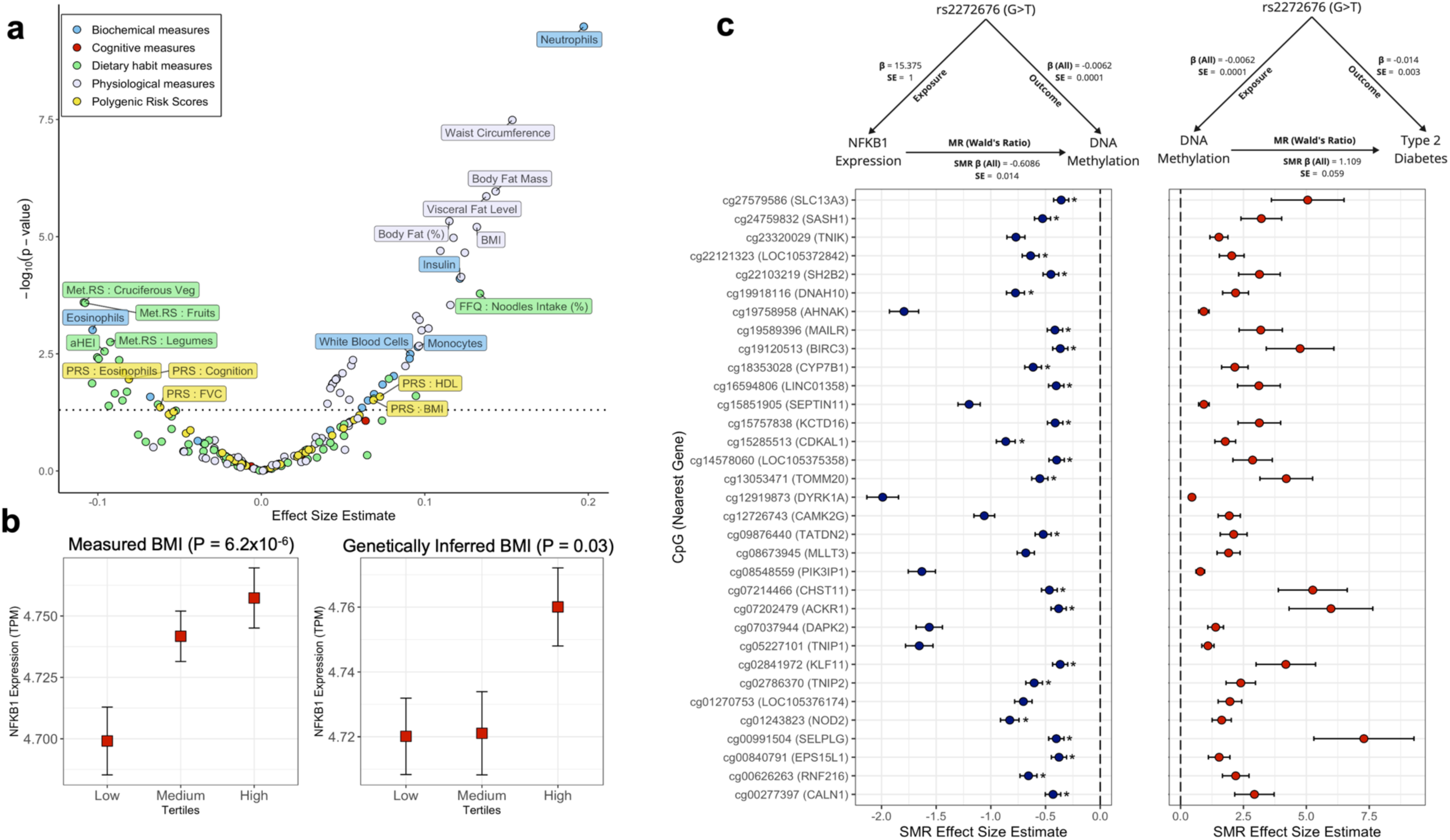
*NFKB1* expression, *trans*-regulation of DNA methylation, and risk of T2D. **Panel a)** Phenome wide association of genomic and epidemiological exposures with *NFKB1* expression. The x-axis represents the regression effect size per SD change in exposure and the y-axis is the -log10(P) for association. Each dot represents an independent phenotype and is coloured by general category. **b)** Box plot showing the range of *NFKB1* expression across the three tertiles of BMI levels and genetically inferred BMI score. P value for association is from linear regression analysis of *NFKB1* expression with the respective phenotype. **c)** SMR analysis between *NFKB1* expression and DNA methylation at sentinel CpGs; and between the CpGs and Type 2 Diabetes using rs2272676 as the instrument variable. CpGs marked (*) showed a shared causal variant with the phenotype (Coloc. PP.H4>0.9); unmarked CpG sites also colocalized but different causal variants (Coloc.PP.H3>0.9). The triangles on top show the direction of SMR analysis. The β_all_ estimate is the fixed effect meta-analysis effect size for all the sentinel CpGs together to show the combined average effect across all associated sentinel CpGs.

### Co-ordinated disturbances of FADS1/2, SREBF1 and related lipid metabolic genes precede the development of diabetes

Our analysis of pleiotropic genetic loci identifies a cluster of 3 *trans*-acting SNPs, which jointly impact 5 diabetes-related CpG loci in *trans* (*ABCG1, SREBF1, MSMO1, DHCR24 and OLMALIN*C; P=2.2x10^-8^ to 3.7x10^-24^; **Figure 4a, Supplementary Table 21**). Increased methylation at the *ABCG1* and *SREBF1* loci shows the strongest relationship to future T2D (RR: 1.18 and 1.14 per 1% change in methylation, P=9.1x10^-46^ and 4.0x10^-32^ respectively; **Supplementary Table 2**), highlighting the potential biological importance of these regulatory connections.

**Figure 4.**
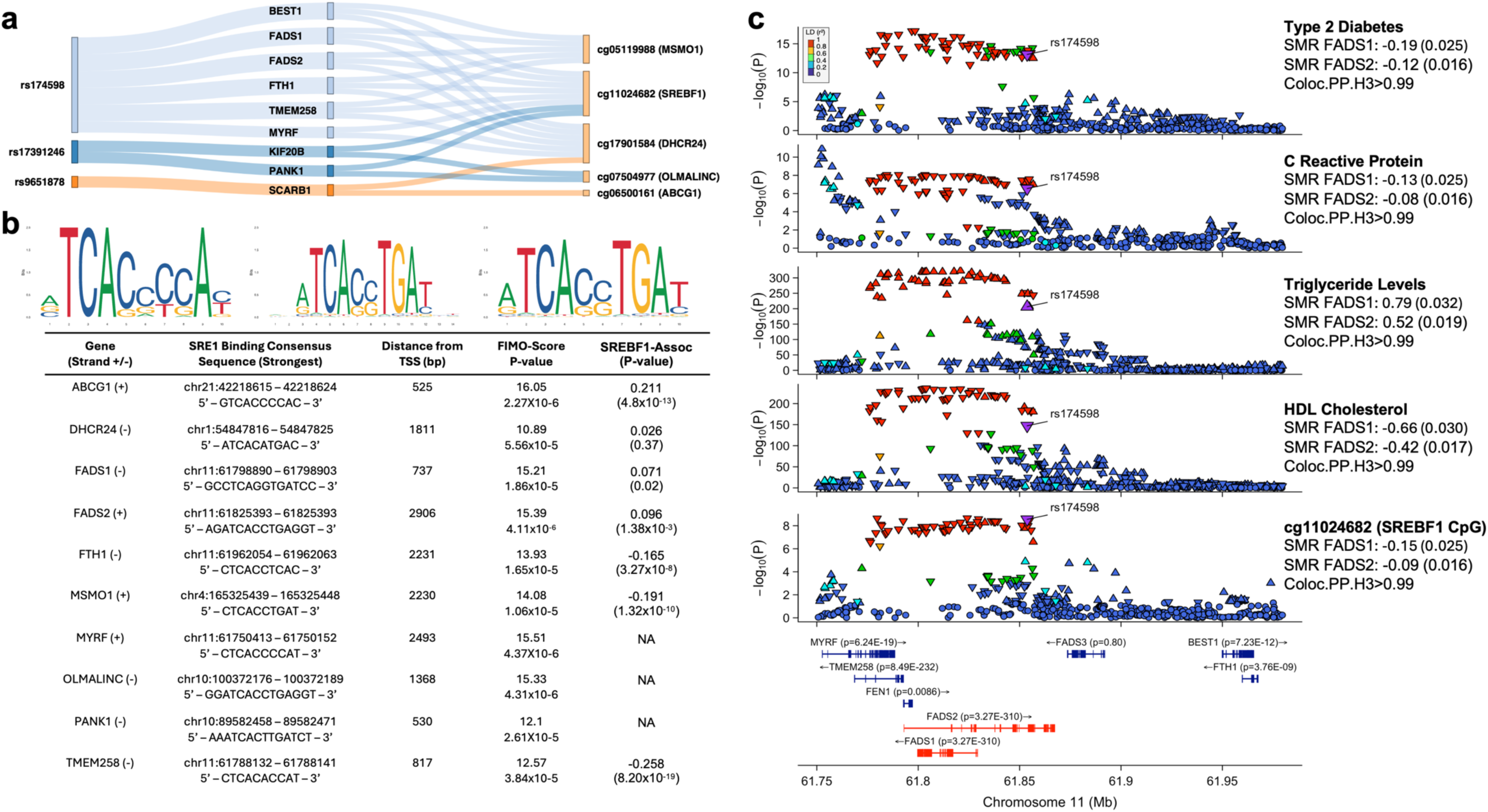
Lipid metabolic gene pathways identified by Sentinel T2D CpGs. **Panel a)** Sankey plot showing the relationships between the core metabolic cluster comprising three *trans-*acting mQTL SNPs, their nine associated cis-eQTLs and five Sentinel CpGs sites associated with T2D. **b)** Sequence motifs of the three known SREBF1 binding consensus sequences obtained from the JASPAR database. The table below highlights the genes from this cluster that have SREBF1 binding sequence in their promoter region, and the association between expression of the named gene and *SREBF1* expression. **c)** Regional plots showing the association of the lead *trans-*acting mQTL SNP (rs174598) at the *FADS1/2* locus with cg11024682 methylation, HDL Cholesterol, Triglyceride levels, C reactive protein and Type 2 Diabetes. The direction of the triangles shows the direction of effect for each SNP on the trait. The colours indicate the strength of LD correlation with the lead SNP. SMR analysis was performed with the lead SNP as the genetic instrument variable, *FADS1* and *FADS2* expression as the exposures and the associated methylation and phenotypes as outcomes.

Amongst the 3 *trans*-acting SNPs in this cluster, rs174598 (GàA) is associated in *cis*, with increased expression of *FADS1* and *FADS2* (eQTLGen Z=40.0 and 62.9 respectively, both P<3.2x10^-3^^20^), encoding key enzymes in fatty acid desaturation and elongation. In keeping with this, SNP rs174598 impacts the concentrations of more than 100 distinct fatty acid metabolites in our participants (P=5.0x10^-8^ to 1.5x10^-208^; **Supplementary Table 23**). SNP rs174598 (GàA) is also associated in *trans* with reduced methylation of cg11024682, the Sentinel CpG at the *SREBF1* locus (Beta=-0.002, P=2.8x10^-9^; **Supplementary Table 21**), and *increased* expression of *SREBF1* (LD proxy SNP rs174577 CàA: *trans*-eQTLGen Z=5.2, P=1.8x10^-7^; LD with rs174598: R^2^=0.86, D’=0.95). SNP rs174598 is also associated with raised triglycerides, lower HDL cholesterol and a lower risk of T2D. SMR and colocalization support a shared genetic basis between *FADS1/2* gene expression, *SREBF1* expression and these key metabolic outcomes (**Supplementary Table 23, Figure 4c**). SREBF1 binds Steroid Regulatory Elements (SRE) to regulate transcription of genes involved in fatty acid synthesis and lipogenesis^35^. Cis-eQTLs that increase SREBF1 expression are associated with raised triglycerides and a *lower* risk of diabetes^28,31^.

Increased methylation of cg06500161 at *ABCG1* is associated with reduced expression of ABCG1 and an increased risk of T2D, suggesting that *ABCG1* activation may reduce risk of T2D. We show an SRE site in the *ABCG1* promoter, and that *SREBF1* and *ABCG1* expression are positively correlated (P=4.8x10^-13^, **Figure 4b**), thus also providing a functional link between these two key metabolic, diabetes predicting methylation loci.

The cluster also identifies *PANK1* and *SCARB1* as potential mediators of *trans-*acting regulation of our diabetes-predicting CpGs (**Figure 4a, Supplementary Table 15**). PANK1 catalyzes the first and rate-limiting enzymatic reaction in the biosynthesis of CoA from vitamin B5, a co-enzyme that plays an essential role in fatty acid metabolism, and in the citric acid cycle^36^. SMR and Colocalization analyses indicate that SNP rs17391246 (AàG) at *PANK1* impacts methylation at both the *SREBF1* and *OLMALINC* loci (*SREBF1*: SMR Beta=0.89, P=1.7x10^-5^; *OLMALINC:* SMR Beta=0.86, P=2.3x10^-5^, both Coloc PP.H3>0.99; **Supplementary Table 21; Supplementary** Figure 6). OLMALINC is a long non-coding RNA that regulates expression of *SCD*, a major gene in lipid biosynthesis, through regional chromosomal DNA-DNA looping interactions^37^. SCARB1 is an integral membrane protein well known to function as the receptor for HDL cholesterol, enabling reverse cholesterol transport^38^. Increased *SCARB1* expression shows genetic colocalization with differential methylation at the *ABCG1* and *DHCR24* loci (*ABCG1*: SMR Beta=-0.35, P=2.55x10^-15^; *DHCR24*: SMR Beta=0.42, P=9.5x10^-21^; both Coloc.PP.H3>0.99; **Supplementary Table 21; Supplementary** Figure 7), as well as with HDL (SMR Beta = - 0.09, P=5.73x10^-34^, Coloc.PP.H3>0.99) and total cholesterol levels (SMR Beta = -0.04, P=3.48x10^-8^, Coloc.PP.H3>0.99, **Supplementary** Figure 7). DHCR24 is an enzyme in terminal cholesterol synthesis, and is linked to vascular disease and cognitive decline^39^.

### Fine-mapping of methylation loci associated with T2D

The MethylEpic array used for our discovery experiment assays only 2-3% of the ∼30M CpG sites in the epigenome^40^. This limits insights into both risk prediction and mechanism. To expand on these array-based association signals, we first carried out whole-genome methylation sequencing (30x depth) of genomic DNA from whole blood, using samples from 500 of our Asian participants (**Extended Data Figure 8**, **Methods**). We used the data to quantify local correlation around our Sentinel CpGs, and to selective genomic intervals for targeted high-depth methylation sequencing.

We show that pairwise correlation >|0.2| with the discovery marker is restricted to <500bp at the majority of CpGs (67.6% of 411 successfully sequenced, **Extended Data Figure 8**). Based on this, we selected a minimum of 500bp either side of the 420 CpGs associated with T2D, for targeted methylation sequencing. At 133 of the methylation loci, there was evidence for correlation at |r|>0.2 beyond the minimum 500bp interval; for these loci we extended the sequencing interval up to a maximum of 5kb in an effort to fully capture this longer range correlation (see **Methods; Supplementary Table 24**). Our primary strategy for methylation sequencing thus aimed to capture DNA methylation at 13,338 unique CpGs (mean 36 per locus, range 1 to 384; **Supplementary Table 24**). As secondary approach, we carried out methylation sequencing for the complete gene region for five methylation loci based on the following criteria: i. Complete resequencing of the *ABCG1* and the *SREBF1* gene locus, as the genomic regions most closely associated with T2D; and ii. three loci with genetic evidence for potential causality of methylation on T2D (*BAIAP2*, *H1-10* and *JARID2*).

High depth (mean coverage: 351x; SD: 92x) targeted methylation sequencing was carried out using a custom sequence capture panel (Twist Bioscience, **Methods**), using genomic DNA from 1,974 whole blood samples (978 cases with incident T2D, and 996 controls). The choice of sequencing depth was based on our previous data, which demonstrates this sequencing depth is needed to achieve precision equivalent to a methylation array^41^. We determined the appropriate statistical threshold for our primary analysis of the relationship between DNA methylation and T2D as P<10^-5^, based on 13,338 independent tests (median ∼14 per locus). Our targeted sequencing had 80% power to identify a difference in methylation between cases and controls of 0.9% at P<0.05, and 1.6% at P<3.7x10^-6^ (Bonferroni correction for 13,338 independent tests), effect sizes that were observed at 56 and 4 respectively, of the 323 Sentinel CpGs identified in the array-based discovery experiment.

### Fine mapping reveals multiple additional methylation signals for T2D

Amongst the 13,338 unique markers assayed in the primary target regions, there was strong evidence for enrichment for association with T2D (λ=1.83). We find 2,252 CpGs associated with T2D at P<0.05, of which 165 CpGs (mean 3, range 1-13 CpGs per locus) are associated with T2D at P<3.7x10^-6^. At 195 of the 314 loci with multiple CpGs captured, the most closely associated CpG was not the marker identified by the array (P_het_<0.05 at 36 loci, **Supplementary Table 25**). Resequencing of the primary intervals thus identifies additional CpG sites associated with T2D, including multiple CpG sites that are more strongly predictive than evaluation based on low-coverage microarray.

Resequencing of the *ABCG1* locus (1MB, 11 genes, 19,216 CpG sites) identified 1,395 CpG sites predicting T2D at P<0.05, and 21 at P<2.6x10^-6^ (**Figure 5**, **Supplementary Table 26**). The methylation markers associated with T2D pile up almost exclusively in the *ABCG1* genic region (20 from 21 [95%] at P<2.6x10^-6^, particularly in *ABCG1* enhancer regions across multiple cell types (FE:3.5 to 16.5, P_FDR_<0.05, 17 cell types; **Extended Data Figure 9**). Of the 21 CpGs at *ABCG1* associated with T2D, 19 were located in introns 1 and 2, in or near annotated LXR response elements (LXRE). The CpGs were most strongly enriched for location in the binding sites for ZNF93 (P=2x10^-11^), a transcriptional repressor, and HAND2 (P=2x10^-9^), a glucocorticoid-responsive TF associated with BMI and implicated in adipogenesis^42^.

**Figure 5.**
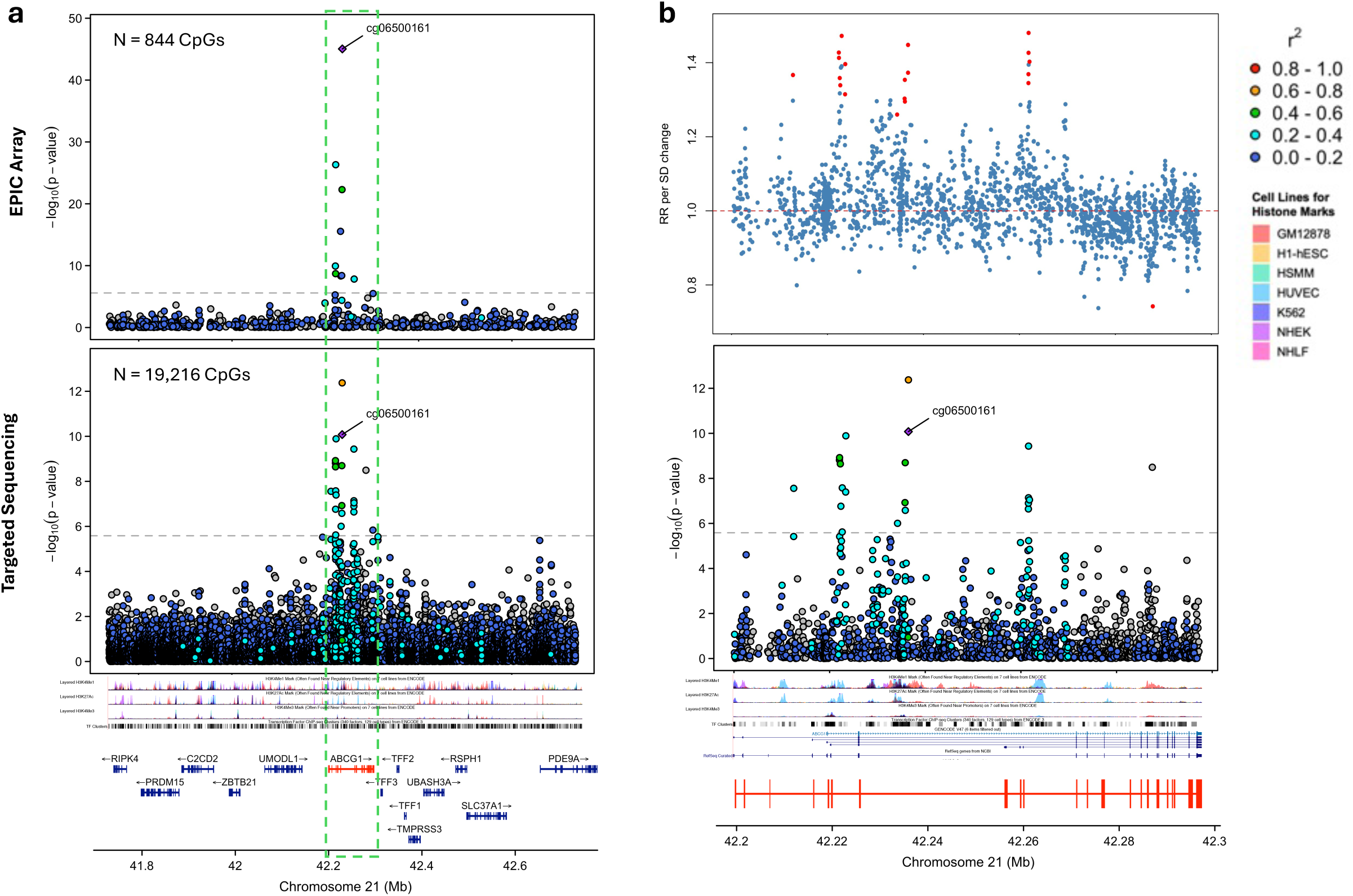
Fine-mapping at *ABCG1* locus. **Panel a)** Top and bottom panel shows the -log10(p) value of association with T2D for CpG sites captured by the EPIC array and TWIST targeted sequencing respectively within a 1Mb region around the sentinel CpG (cg06500161). **b)** Zoomed in regional plot of *ABCG1* genic region, as indicated green rectangle in. Top panel shows the relative risk (RR) for T2D per standard deviation (SD) change in methylation level, whilst the lower panel indicates the -log10(P) value of association with T2D for CpG sites within the *ABCG1* genic region. Information about the regulatory regions was obtained from UCSC genome browser for seven cell lines and are highlighted and labelled in the legend. (GM12878: Lymphoblastoid cells; H1-hESC: H1 human embryonic stem cell line; HSMM: Human skeletal muscle myoblasts; HUVEC: Human umbilical vein endothelial cells; K562: human chronic myelogenous leukemia (CML) cell line; NHEK: Normal Human Epidermal Keratinocytes; NHLF: Normal human Lung Fibroblasts)

The most closely associated CpG in *ABCG1* was now located at chr21:43656590 (RR [95% CI]:1.44 [1.31 to1.60] per SD; P=1.1x10^-12^), a marker not assessed by the methylation arrays. Furthermore, a gene-based methylation score (developed using least absolute shrinkage and selection operator [LASSO] modelling, see **Methods**) that incorporated 17 of the 21 CpGs associated with T2D at *ABCG1*, achieved a RR for T2D of 2.12 per SD increase in methylation score (95% CI: 1.71 to 2.62; P=5.1x10^-12^). This represents a ∼3-fold improvement compared to the effect size for the initially identified array-based Sentinel CpG cg06500161 (RR for T2D: 1.37 [1.25 to 1.51] per SD; P=1.5x10^-10^; P_het_=2.8x10^-4^). Similarly, at the *SREBF1* locus (1MB, 18 genes, 16,282 CpG sites), we identify 1,011 CpG site predicting T2D at P<0.05, and five at P<3.1x10^-6^ (**Supplementary Table 27**). The five T2D- associated CpG sites were again found exclusively in the *SREBF1* gene body (**Extended Data Figure 10**) and were also enriched in enhancer regions (FE:2.4 to 11.5, P_FDR_<0.05, 30 cell types; **Extended Data Figure 9)**. As was the case for the *ABCG1* locus, the gene-based approach at the *SREBF1* locus also resulted in an improved relative risk, with a RR for T2D of 1.93 (1.50 to 2.48) per SD (P=2.7x10^-7^), representative of a 58% improvement compared to the initially identified array-based sentinel CpG (P_het_=0.01). Fine-mapping of the methylation markers identified by the microarray thus offers new insights into genomic regulation, and identifies multiple novel CpG associated with T2D, that as a set strongly predict future T2D.

Amongst the three loci selected for fine-mapping based on evidence of a potential causal relationship between methylation and T2D from colocalization, at *BAIAP2* and *H1-10* loci we found only one CpG associated with T2D at P<0.05 after Bonferroni correction for the number of CpGs sequenced. At the *JARID2* locus, none of the CpGs achieved statistical significance.

### Potential clinical relevance: Methylation Risk Scores predict future diabetes

As a key objective, and building on our prior work^4,43^, we tested the potential utility of DNA methylation for prediction of T2D. Having identified multiple predictive methylation markers using our array-based analyses, we first used a LASSO model to identify a parsimonious set of 42 array-based CpGs that independently predict T2D, amongst participants of the MEC study (‘Development set’, **Supplementary Table 28**). We then estimated an epigenome-wide Methylation Risk Score (MRS) as the sum of the site-specific DNA methylation levels, weighted by effect size for association with T2D, for each of the 42 independent CpGs. We tested the MRS for prediction of T2D amongst participants of the iHealth-T2D study (‘Validation set’). We compared effect sizes and discrimination of future T2D for the MRS, and for a diabetes polygenic risk score (PRS). Based on our prior work, we carried out a pre-specified analysis of MRS for prediction of T2D amongst normoglycaemic and obese individuals; this approach recognises that there is an important need to improve prediction of diabetes amongst obese people before the onset of prediabetes.

Our array-based MRS is strongly predictive for future T2D in the validation dataset (RR: 5.4 [3.9 to 7.5] in Q4 vs Q1 of MRS, P=6.3x10^-24^; **Figure 6**; **Supplementary Table 29**), MRS is as predictive for future T2D as established PRS scores (RR: 4.7 [3.4 to 6.4] in Q4 vs Q1 of PRS, P=2.3x10^-22^), with similar discrimination (AUC: 0.7 vs 0.65 respectively, P=0.13). The relationship of MRS and PRS with T2D were independent and additive, consistent with the view that MRS identifies additional, non-genetic, behavioural exposures underpinning development of diabetes (**Supplementary Table 29**). MRS remained predictive for T2D after adjusting for BMI, glucose and HbA1c concentrations (RR: 2.5 [1.7 to 3.8] in Q4 vs Q1 of MRS, P=3.0x10^-6^). MRS also remained strongly predictive for future T2D amongst key subgroups, including the subset of normoglycaemic people with overweight and obesity (RR: 5.0 [3.0 to 8.1] in Q4 vs Q1 of MRS, P=1.8x10^-10^; (**Supplementary Table 29**). The array-based MRS also replicated in the 2,664 South Asians from the LOLIPOP study, despite the reduced number of available CpG sites (RR: 6.8 [5.1, 8.9] in Q4 vs Q1 of MRS, P=1.1x10^-41^), and in a fully independent cohort of 255 East Asians in the SCHS study (RR: 8.0 (2.8 to 22.7) in Q4 vs Q1 of MRS, P=9.9x10^-5^; **Supplementary Table 29**). The relationships of MRS with T2D in the replication were also independent of BMI and measured glycaemic risk factors.

**Figure 6.**
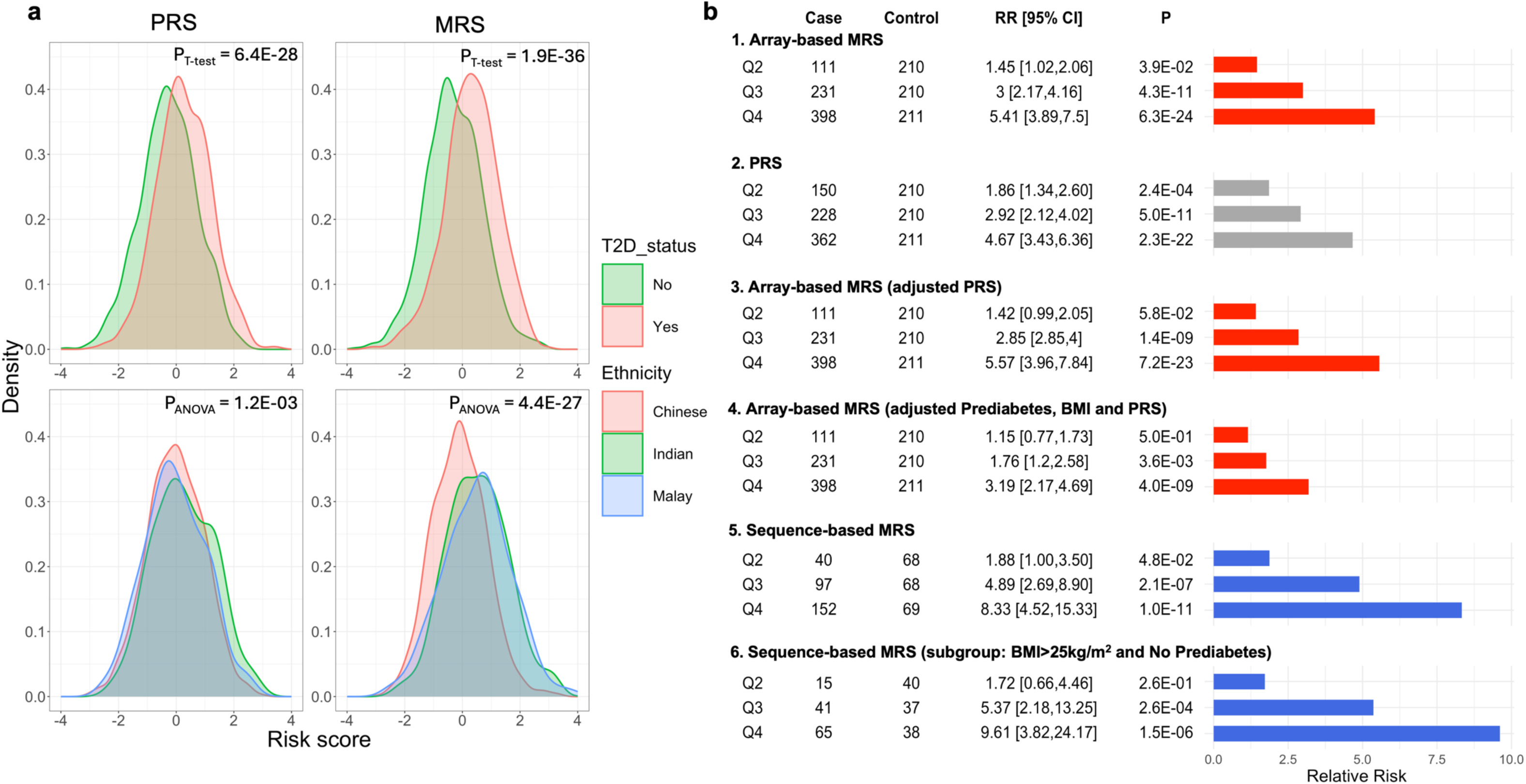
Methylation Risk Scores and Type-2 Diabetes in Asian populations. **Panel a).** Density plot showing the distribution of MRS and PRS values in T2D cases and controls (upper two quadrants) and between Asian ethnic subgroups (lower two quadrants) in the HELIOS study. **b)**. Association of Array-based and Sequence-based MRS with T2D. Model 1: Array-based MRS and T2D in the 1,663 iHealth-T2D participants (‘Test set’), compared to PRS (Model 2), after adjustment for PRS (Model 3) and additional adjustment for Prediabetes (Model 4). Model 5: Sequence-based MRS and T2D in 588 samples from the targeted sequencing experiment, used as the ‘Test set’. Model 6: Sequence-based MRS and T2D in the subset of participants with overweight or obesity (BMI>25kg/m2) but without Prediabetes. Samples were separated into quartiles based on distribution in the controls. Associations with T2D was tested by logistic regression. Relative Risks for T2D are reported relative to Quartile 1 (reference). All analyses are adjusted for age and sex.

We hypothesised that CpG sites assessed through resequencing would enable us to improve on the array-based MRS. We applied LASSO modelling to the sequence dataset, starting with the set of 165 CpG sites captured by fine-mapping that were associated with T2D at P<3.7x10^-6^ **(Supplementary Table 31)** The sequence-based MRS based on 60 LASSO-selected CpGs, which represents a 2.2-fold improvement over our array-based MRS (RR for T2D per 1SD increase in MRS: 2.59 [2.07 to 3.25], P=7.7x10^-17^ vs 1.73 [1.56 to 1.91], P=1.9x10^-27^; P_het_=0.001) and for Q4 vs Q1 of MRS (RR: 8.3 [4.5 to 15.3], P=1.0x10^-11^ vs 3.9 [3.0 to 5.2], P=1.6x10^-22^ in the same set of samples; P_het_<0.05). (**Supplementary Table 32; Supplementary Table 33**). Our sequence-based MRS remains strongly predictive for future T2D after adjusting for prediabetes (RR in Q4 vs Q1: 5.8 [3.0 to 11.3], P=2.4x10^-7^), and was highly discriminatory amongst a prespecified group of normoglycaemic individuals with overweight or obesity achieving a RR for T2D of **9.6 [3.8 - 24.2]** between Q4 and Q1 (P=1.5x10^-6^). AUCs for prediction of diabetes were similar for MRS and prediabetes as separate exposures (0.72 [0.68 to 0.76] vs 0.72 [0.68 to 0.76] respectively, P=0.98). The addition of the sequence-based MRS to prediabetes as a risk factor improved prediction significantly (AUC for T2D: 0.80 [0.77 to 0.84], P=4.8x10^-6^ compared to Prediabetes alone). The prospects for clinically useful risk stratification are thus strengthened by sequencing-based approaches, which show improved discrimination compared to array-based MRS.

### Methylation Risk Scores vary between ethnic groups

The incidence and prevalence of T2D are recognised to vary greatly both between Asian ethnic groups, and amongst Asians compared to Europeans^1,3–5^. Similarly, we show that there is a 4.5 fold higher risk of diabetes amongst South Asians, compared to people of Chinese ancestry, amongst our Asian participants (**Supplementary Table 34**). We find that MRS varies between Asian ethnic groups in close relationship to their different metabolic risk (P=4.13x10^-40^; **Figure 6**). Array-based MRS is elevated amongst South Asians and Malays compared to Chinese (mean: 0.52 and 0.50 vs -0.10 AU respectively; P=1.2x10^-18^) and explains 51% of the 4.5 fold excess risk for T2D in South Asians, and 67% of the 3.1 fold excess risk for T2D in Malays. In contrast, PRS for T2D are similar between Asian ethnic groups, and explains just 13% of the excess risk of T2D amongst South Asians, and only 1% of the excess risk In Malays, compared to Chinese. Similarly, array-based MRS is raised in South Asians compared to Europeans living in the UK (LOLIPOP study; 0.02 vs -0.36; P=2.6x10^-6^), and explains ∼24% of the 3.1 fold increased risk of diabetes in Asians. Given the improved discrimination of sequencing-based compared to array-based MRS, we anticipate that sequencing-based MRS are likely to further improve the ability to explain the difference in risk for T2D between populations.

## DISCUSSION

The burden of T2D is high and rising amongst Asian populations, including early-onset, lean and insulin resistant phenotypes, that appear distinct from European metabolic phenotypes^6,7^. Building on previous work, we set out to both understand the nuclear regulatory mechanisms underlying increased susceptibility to diabetes and related metabolic disturbances in Asian populations, and to identify potential biomarkers to advance risk stratification. We focused on exploring DNA methylation at CpG dinucleotides, one of several reversible chemical modifications to the structure of DNA, which contributes to both physiological regulation of gene expression, as well as the perturbations in genome regulation that contribute to developmental disorders, senescence, cancer and other chronic diseases^44^.

### DNA methylation loci associated with future diabetes in Asian populations

We carried out epigenome-wide association in peripheral blood white cells, from three longitudinal cohorts, comprising Asian people well-characterised for incident T2D. Methylation profiles vary by cell lineage^44^, and our observations thus relate primarily to regulatory disturbances in blood cells. Nevertheless, our choice of tissue was motivated both by the close involvement of white cells in the aetiology of insulin resistance, inflammation, lipid metabolism and the vascular consequences of diabetes, as well as by the suitability of blood cells for clinically applicable biomarker discovery.

We found 323 Sentinel CpGs from 314 loci associated with future T2D. We replicate all previously reported associations of DNA methylation in whole blood with future T2D, including the *ABCG1*, *SREBF1* and *TXNIP* loci^4,10–14^, but also identify more than 300 novel methylation loci predictive for diabetes. Effect sizes of the methylation sites for T2D risk are higher than for common genetic variants and are consistent across both cohorts and Asian ethnic subgroups. We note that perturbed methylation at our Sentinel CpGs identifies both genetic and environmental exposures known to be involved in the aetiology of diabetes, including adiposity, educational attainment, and unfavourable dietary intake. This not only lends plausibility to the involvement of the methylation loci in the pathways to diabetes, but also highlights the CpGs as potential biomarkers to identify multifactorial exposures influencing metabolic health.

We used *cis-*acting genetic instruments, and colocalization analyses, to show that DNA methylation at the *BAIAP2*, *CPNE6*, *H1-10*, *HMGA1, INAFM2*, *JARID2, MDM4*, *RPS6KA2*, *SLC12A2* and *TGM4* loci may be causally linked to the development of T2D, and that multiple methylation loci may share a common genetic basis with adiposity, lipid metabolism and cardiovascular risk. BAIAP2 functions as an insulin receptor tyrosine kinase substrate, and may be involved in insulin signalling in the central nervous system, and hypothalamic pathways for regulation of adiposity^45^. *INAFM2* encodes InaF-motif containing 2, a protein of unknown function. Genetic variants at *INAFM2* are associated with T2D in Asian, but not European populations^46,47^. MDM4 regulates p53 during embryonic development and adulthood in a cell and tissue-specific manner, and is required for development of endocrine pancreas. *MDM4*^-/-^ mice show inhibition of islet cell proliferation, and pancreatic beta cell dysfunction^48^. *JARID2* encodes a DNA binding transcriptional repressor with a key role in the development of the endocrine pancreas^49^. *Jarid2*^-/-^ mice have reduced beta-cell mass, impaired insulin secretion and glucose intolerance^49^. *SLC12A2* encodes an ion transporter expressed in pancreatic β-cells. Mouse models with an *Slc12a2^-/-^* knockout have reduced in vitro insulin responses to glucose and islet hypoplasia, and demonstrate weight gain and a progressive metabolic syndrome phenotype^50^. The methylation loci thus identify *cis*-candidate genes, with compelling evidence for a role in metabolic health.

### DNA methylation patterns identify disturbances of immune and metabolic control

Multiple lines of evidence suggest that the Sentinel CpG identify functionally relevant genomic regions, including *trans-*acting nuclear pathways. We took advantage of additional multi-ethnic Asian population studies, with rich and molecular phenotyping to understand the exposures, genes and biological processes involved. Using *trans*-acting genetic instruments, we show that the Sentinel CpGs are components of transcription factor pathways that regulate key co-ordinated inflammatory, lipid and glucose metabolic pathways of T2D. In particular, the CpGs intersect, and are causally influenced by *NKFB1, NFKBIA, NFKBIE, NF1A, COMMD7, IKZF3, MADD* and other nuclear transcription factors involved in immune regulation and metabolic control. Immune activation is a well-recognised manifestation of increased adiposity and diabetes, and is reported to contribute to pancreatic beta cell dysfunction, insulin resistance, atherosclerosis and steato-heptatitis^51^. We show that both measured adiposity and genetically inferred adiposity are associated with *NFKB1* activation, which correlates closely with methylation at multiple Sentinel T2D CpG sites in *trans.* Interestingly, our genetic instrument analyses confirm causal association between *NFKB1* expression and DNA methylation, but also suggests that the *NFKB1* activation may reduce the risk of T2D. Some aspects of immune activation may thus operate to mitigate against the increased risk for diabetes characteristic for excess adiposity.

The *trans*-acting networks also include *CDKAL1, CPT1A, CYP7B1, PDK4, LDLRAD2, SREBF1, SH2B2, SOCS3, TANK* and *TXNIP*, genes reported to impact pancreatic beta cell function, insulin signalling and action, glucose sensing, metabolism of glucose, cholesterol and lipids, fatty acid beta oxidation, mitochondrial biology, thermogenesis, and adipogenesis^52–59^. Using genetic variants as instrumental variables, we further show that these *trans*-acting pathways, and their associated CpGs are causally linked to diabetes, as well as to closely related phenotypic disturbances such as adiposity, lipid levels, systemic inflammation, and cardiovascular disease. Our Sentinel CpGs thus identify both genes and pathways contributing to the pathogenesis of diabetes and its related cardiovascular and metabolic abnormalities.

### Disturbances in ABCG1 and SREBF1 as early markers for T2D susceptibility

DNA methylation at the *ABCG1* and *SREBF1* loci showed the strongest relationship to future T2D in both our array and targeted sequencing datasets, with increased methylation associated with *lower* expression of the respective genes in white cells, and a *higher* risk of T2D. Increased methylation at *ABCG1* and *SREBF1* is also linked to obesity, insulin resistance, atherogenic dyslipidaemia and cardiovascular risk, as well as to the unfavourable dietary habits that drive these conditions^43,60–63^. Suppression of *ABCG1* and *SREBF1* gene expression thus appears to contribute to risk for diabetes and related metabolic disturbances.

ABCG1 is an ATP-binding cassette (ABC) transporter, and a critical component the cholesterol and phospholipids from cells, including reverse cholesterol transport^64^. *Abcg1*-/- mice have impaired cholesterol efflux from macrophages, resulting in the accumulation of cholesterol in tissues, disruption of insulin release by pancreatic beta cells, foam cell formation and atherosclerosis^65,66^. SREBF1 is recognised to be a key regulator of lipid metabolism, which induces both fatty acid biosynthesis and de novo lipogenesis in the liver and other tissues, through binding to SRE^67,68^. In addition, knockout and over-expression studies show that increased expression of *SREBF1* in the pancreas promotes β-cell proliferation and insulin secretion, providing a potential compensatory mechanism to metabolic stress^69^, potentially mitigating against the lipo-toxicity and the development of pancreatic β-cell failure, which underpins the transition to diabetes. Our pathway analyses using *trans*-acting genetic instruments further link *ABCG1* and *SREBF1,* and other genes also documented to play key roles in fatty acid and lipid metabolism, including *FADS1, FADS2, SCARB1, DHCR24 and OMALINC*^37–39^. Our findings thus identify aberrant lipid metabolism and transport as early, predictive and predominantly environmentally-driven phenotypes, in the development of diabetes in Asian populations.

### Fine mapping of the T2D associated methylation loci

The MethylEpic array assays just 3% of genomic CpG sites^40^; this sparse coverage limits insights into biological mechanisms and clinical utility for risk prediction. To move beyond state-of-the-art, we carried out fine-mapping of T2D associated CpGs, using a combination of whole-genome methylation sequencing to inform correlation structure, and targeted high-depth methylation sequencing to quantify disease associations. These approaches have not previously been deployed in studies of DNA methylation and metabolic disease.

We show that pairwise correlation between markers is typically short range (<500bp), in contrast to the long-range LD that is well established for DNA sequence variation. Complete resequencing of the *ABCG1* and *SREBF1* loci revealed multiple CpGs that associate with T2D, and located almost exclusively in intronic enhancer regions of the respective candidate genes. At other loci, we also find multiple additional, and previously uncharacterised CpGs associated with T2D. Furthermore, at 195 of the 314 loci, the most closely associated CpG was not the marker identified by the array. Resequencing of the primary intervals thus offers new insights into genomic regulation, and identifies additional CpG sites associated with T2D, including multiple CpG sites that are more strongly predictive than evaluation based on low-coverage microarray.

### Prediction of T2D and potential clinical utility

Clinical intervention studies show that T2D can be prevented through lifestyle and pharmacological interventions. As a result, there is considerable interest in accurate identification of people at risk for T2D, to enable them to be offered effective therapeutic interventions. Current approaches to risk stratification for T2D rely heavily on markers of disturbed glucose metabolism, including impaired fasting or post-load glucose concentrations, or an elevated HbA1c. Whilst these markers do identify people with a high risk for T2D, abnormal glucose metabolism represents a late stage in the progression to diabetes. Risk of both micro-vascular and macrovascular disease is already increased amongst people with impaired glucose handling^70^.

To address the need for biomarkers that improve prediction of T2D, in particular amongst people with normal glucose concentrations, we tested the potential utility of DNA methylation for prediction of T2D. We show that a methylation risk score, comprising a parsimonious set of 42 array-based CpGs, is strongly predictive for future T2D, with similar discrimination to polygenic information. MRS remained predictive after adjusting for BMI, glucose and HbA1c concentrations, and remained predictive for future T2D amongst the subset of normoglycaemic people with overweight and obesity. Incorporation of information from targeted sequencing further improved model performance. Specifically, our sequence-based MRS, based on 60 CpGs, achieved a 2.2-fold improvement in risk ratio compared to our array-based methylation score. The potential importance of MRS for risk stratification is further illustrated by comparison between our Asian ethnic subgroups. In particular, we show that MRS explains much of the difference in risk for T2D between people Asian subgroups, and an important component of the difference between South Asians and Europeans. In contrast, polygenic information provides little explanation for the differences in metabolic outcomes between Asian populations. Our findings thus strengthen the basis for the potential utility of DNA methylation to identify individuals at high-risk of T2D for preventative interventions, including amongst people in the earliest stages of the progression to diabetes who have not yet developed impaired glucose metabolism.

### Summary

We found disturbances in DNA methylation at hundreds of CpG sites in peripheral blood white cells that predict future T2D. These highlight a critical role for nuclear regulatory disturbances, including in both immune activation pathways, and metabolic networks involving *FADS1/2*, *SREBF1*, *ABCG1* and other genes causally implicated in pancreatic beta cell function, insulin signalling, fatty acid metabolism and lipid transport. The CpGs are highly predictive for future T2D, independent of adiposity and glycaemic risk factors, and additive to genetic risk. Our research thus provides new insights into the regulatory disturbances that precede development of T2D, and reveals the potential for DNA methylation biomarkers to identify genetic and non-genetic exposures influencing diabetes, and inform risk stratification in diabetes prevention.

## METHODS

### Population samples description

### Multi-Ethnic Cohort (MEC)

MEC is a prospective population study of men and women of Chinese, Indian or Malay ethnicity, aged 21 to 85 years, and living in Singapore. Participants were recruited from the general population, between 2004 to 2010^15^. At enrolment all participants completed a detailed interview, physical examination, and provided blood samples at each visit. Anthropometric measures such as height, weight, waist and hip circumferences were measured in the physical examination. T2D status, age, gender, smoking and alcohol drinking patterns, medication history and other covariates were collected through detailed interview. All participants provided written formed consent, and all protocols associated with the study were approved by the National University of Singapore Institutional Review Board.

### The iHealth-T2D study

iHealth-T2D is a prospective study of Indian Asian men and women living in West London, and recruited in 2016. At enrolment all participants completed a structured assessment of cardiovascular and metabolic health, including anthropometry, and collection of blood samples for measurement of fasting glucose, lipid profile and HbA1c. Identification of incident T2D was done through face-face follow-up, supplemented by linkage to medical records. Epigenome-wide association was performed using genomic DNA from peripheral blood collected at enrolment. The study is approved by the National Research Ethics Service (16/WM/0171) and all participants gave written informed consent.

### The London Life Sciences Prospective Population Study (LOLIPOP)

LOLIPOP is a prospective cohort study of ∼28K Indian Asian and European men and women aged 35 to 75 years, recruited from the lists of 58 General Practitioners in West London, United Kingdom between 2003 and 2008. At enrolment all participants completed a structured assessment of cardiovascular and metabolic health, including anthropometry, and collection of blood samples for measurement of fasting glucose, insulin and lipid profile, HbA1c, and complete blood count with differential white cell count. Aliquots of whole blood were stored at -80C for extraction of genomic DNA. Epigenome-wide association was performed using genomic DNA from peripheral blood collected at enrolment, as previously reported^4^. The LOLIPOP study is approved by the National Research Ethics Service (07/H0712/150) and all participants gave written informed consent.

### Health for Life in Singapore (HELIOS)

The HELIOS study recruits Singaporean citizens and permanent residents aged 30 to 84 years old from the general population^17^. Study participants complete questionnaires on demographic and medical information, have measurements across various system domains including anthropometry, and collection of blood samples for measurement of fasting glucose, insulin and lipid profile, HbA1c, and complete blood count with differential white cell count. Aliquots of whole blood were stored at -80C for extraction of genomic DNA. Epigenome-wide association was performed using genomic DNA from peripheral blood collected at enrolment. The HELIOS study is approved by the Nanyang Technological University Institutional Review Board (IRB-2016-011-030), and all participants gave written informed consent.

### Singapore Chinese Health Study (SCHS)

The Singapore Chinese Health Study (SCHS) is a prospective cohort study of 63,257 Chinese men and women aged 45 to 74 years, residing in Singapore, and recruited between 1993 and 1998 as previously described^71^. At baseline, all participants completed structured interviews capturing demographic data, dietary patterns, lifestyle behaviours, and medical history. Anthropometric measurements, including height and weight, were recorded, in two subsequent follow-ups carried out in 1999-2004 (Follow-up I) and 2006-10 (Follow-up II), which also included collection of blood samples from a subset of participants. The study was ethically approved by the relevant Institutional Review Boards, and informed consent was obtained from all participants.

Within the SCHS cohort, a nested case-control study for diabetes was completed, as previously described^72^. In brief, we this comprised participants who were free of diabetes, cardiovascular disease, and cancer both at baseline and Follow-up I. Cases included participants diagnosed with diabetes during Follow-up II (2006-2010). Controls were selected from those free of the specified diseases at Follow-up II, matched on age, date of blood collection, sex, and dialect group. The present study included 255 cases of incident diabetes and controls, who had samples of whole blood available from the first follow-up evaluation.

### SG10K_Health

The SG10K_Health study is part of the Singapore National Precision Medicine programme, and represents a collaboration between five adult population cohorts and one paediatric cohort in Singapore. Participants were of Chinese, Indian and Malay ethnicities and recruited with approval of a relevant institutional ethics review board, as previously described^73^. For the current study, we used the whole genome genotyping and DNA methylation profiles (Illumina HumanMethylationEPIC array) previously generated for the adult participants only (N=7,749).

### Quantification of DNA methylation and quality assessment of samples and markers

DNA methylation was quantified using the Illumina HumanMethylationEPIC array (EPIC array) for the MEC, iHealth-T2D, HELIOS and SCHS samples. Genomic DNA was bisulfite converted using the EZ DNA methylation kit according to manufacturer’s instructions (Zymo Research). Bisulfite converted genomic DNA was quantified using the EPIC array, according to manufacturer’s instructions. Bead intensity was retrieved using *minfi* R package, and downstream analyses are conducted using *minfi* and R^74^. Illumina background correction was applied to all intensity values using *minfi*. Methylation markers on sex chromosomes are excluded. A detection P-value threshold of P<0.01 was used to set intensity values to NA. The proportion of missing data points per sample or per marker were determined, and samples or markers with low call rate (<98%) were excluded. Gender swapped samples identified by *minfi* were also excluded.

A total of 12 samples were excluded for MEC due to gender swaps, leaving 1,492 samples for analysis. No sample in the MEC cohort failed the call rate threshold of 98%. Of the 846,459 autosomal CpG markers assayed by the EPIC array, 3,465, 3,539 and 3,762 markers with call rates <98% were excluded in the MEC Chinese, Malay and Indian subgroups respectively, leaving 842,994, 842,920 and 842,697 markers on the autosomes. In the iHealth-T2D study, 50 samples were excluded for low sample call rate and 12 for gender swaps, leaving 1,663 samples for analyses. 3,446 markers were removed for low marker call rates (<98%), This left 843,013 markers on the autosomes. Processing of samples for the LOLIPOP study has been previously described^4^.

In HELIOS, 837,722 CpG markers were successfully assayed. In total 58 samples were excluded; two for array scanning failure, 39 for gender inconsistency and 17 duplicates. A further 105 samples that were not from the three major Asian ethnic groups, were also excluded. This left us with 2,237 samples for analysis. There were 255 samples successfully assayed on the SCHS study.

### Statistical analyses of epigenome-wide data

Primary analysis of epigenome-wide data was performed as described previously^75^. Briefly, marker intensities were quantile normalised. Normalised intensity values were then used to calculate the beta value (methylation level at each CpG site). Control probes of EPIC array intensities were retrieved using *minfi.* Principle component analysis (PCA) of control probe intensities was performed and the resulting PCs 1 to 30 were included as predictors in the subsequent regression models to adjust for technical biases. The proportion of six white blood cell sub-populations (CD8 T cells, CD4 T cells, Natural Killer cells, B-cells, Monocytes, Granulocytes) were estimated using the Houseman methods^76^. The association of each autosomal CpG site with incident T2D was tested using logistic regression, adjusted for confounders such as age, gender and further adjusted for imputed white blood cells (WBC) proportion and PC1-30 of control probe intensities, as these progressively reduced test statistic inflation.

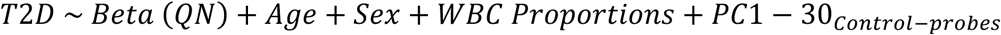

The regression analyses were performed on individual ethnic groups separately for MEC. Results were combined across the three ethnic groups by meta-analysis using METAL^77^. We then performed a second round of meta-analysis to combine results between MEC, iHealth-T2D and LOLIPOP datasets. Epigenome-wide significance was set at *P*<8.62x10^-8^; this threshold was based on the results of permutation testing^16^, and is similar to the threshold that would have been obtained via Bonferroni correction for the 848,166 autosomal markers tested.

### Phenome-wide association analysis

We tested the associations between the 323 Sentinel CpGs (1 sentinel CpG not in the 850K EPIC array), and 185 trait-exposures, including directly measured phenotypes as well as genetically inferred exposures calculated as Polygenic Risk Scores (PRS). The summary statistics of the different traits were obtained from the PGS Catalog^23,24,78–82^ and the PLINK score function^83^ was used to estimate the PRS for the individuals in the HELIOS cohort. We then performed regression analysis for association of methylation with the respective trait, with adjustment for age, sex, ethnicity, methylation array control probe PCs, and proportion of six white blood cell sub-populations estimated by the Houseman method^76^.

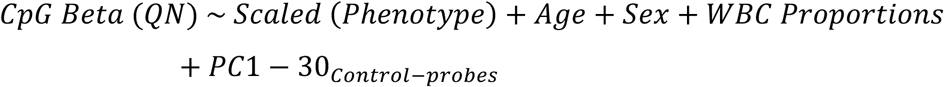

To estimate the enrichment of associations within the sentinel CpG sites compared to background, we repeated the regression analysis for 1000 matched random sets of 323 CpGs. The matching was performed based on mean methylation beta (± 5%), standard deviation (± 0.5%), and presence in methylation array (present in 850k array only or present in both 450k and 850k array). For each trait, we calculated the fold enrichment (FE) as ratio of number of sentinel CpG set (observed) to the mean number of CpGs (1000 random sets, expected) associated with trait at P<0.05, The empirical P value for the FE was obtained based on the distribution of expected (i.e.: P_perm_ = N(Random sets with count≥ Sentinel Count)/1000)

### Functional Annotation of Sentinel CpGs

We performed functional overlap analysis of the sentinel CpGs to evaluate the enrichment of the 323 sentinel CpG sites across DNase I hotspots, 5 histone marks and 15 chromatin states across 39 cell types from the Roadmap Epigenomic Consortium (https://egg2.wustl.edu/roadmap/web_portal/index.html)^84^. We mapped the genomic location of each of the sentinel CpG to understand whether they were present at the different biologically relevant locations in the genome. To determine whether the overlap occurred more often than expected by chance, we determined the total number of CpGs overlapping with the different locations. For each epigenetic mark, we calculated the number of overlapping sites amongst the 323 replicating markers (observed) and all the array based CpGs (expected). We calculated FE as proportion of observed count /proportion of expected. The P-value for enrichment and depletion was estimated using hypergeometric test for over-representation and under-representation respectively, which were then adjusted using FDR to correct for multiple testing.

### Transcription Factor (TF) Enrichment

The binding site information for 1210 human TFs tested was obtained from the Remap database, 2022 release (https://remap.univ-amu.fr/)^85^. We used the Homo sapiens Cis Regulatory Modules (CRM) peaks for this analysis. We first determine how many of our sentinel CpGs overlap with the binding sites of the different TFs, and then estimated FE and the hypergeometric P value for enrichment by comparing the proportion of overlap of our sentinels to the proportion of overlap of all CpG probes. We then applied FDR based P value adjustment to correct for multiple testing.

### Methylation and Gene Expression

Gene expression data was available for 12,434 genes and 1,228 participants from the HELIOS study. Generation and processing of the RNA sequencing data has been previously described^17^. We performed association analysis between DNA methylation at sentinel CpGs and gene expression to identify both *cis-* and *trans-*eQTMs associated with the sentinel CpGs. Analysis was performed using matrixQTL tool^86^ adjusting for Age, Sex, Ethnicity, RIN, top six PEER factors and the six white blood cell proportion estimates.

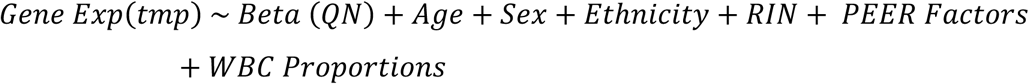

Significant associations were determined after Bonferroni correction for the number of pairwise test performed. For the *cis*-eQTMs, we evaluated the importance of gene proximity by investigating associations under different distance thresholds (nearest gene, genes within 500KB, genes with 1MB). We observed enrichment for association of CpGs only with nearest gene in *cis*. For functional genomic analysis of CpGs in *cis*, including annotation of *cis*-eQTMs, we therefore only consider nearest gene. In our *trans*-eQTM analysis, we tested associations between sentinel CpG and all the quantified *trans*-genes (distance between CpG-Gene >10MB) in the dataset. CpG-Gene pairs with P<0.05/12,434 were considered significant (ie Bonferroni correction for number of genes tested).

To determine if the sentinel CpG sets have a greater number of *cis* and *trans* eQTMs compared to the background, we repeated the eQTM analyses for the 1000 random sets of CpGs, using the same methods. FE and empirical P value for enrichment of both *cis*- and *trans*-eQTM counts amongst sentinel CpG, was determined in comparison to the respective eQTM counts for the random CpG sets.

We investigated whether the sentinel CpGs, and their associated gene expression signatures, show covariation that is greater than under the null expectation (random CpG sets). We calculated all pairwise correlations between the sentinel CpGs to obtain the Observed distribution. We repeated the same in the 1000 randoms sets (Background). We then absolute values for pairwise correlation between Observed and Background, and determined the FE (mean[absolute pairwise correlation in Observed]/ mean[absolute pairwise correlation in Background]) and P value (t-test). Similar analysis was performed for the *cis* and *trans*-eQTM signatures of the sentinel CpGs.

### Gene Set Enrichment Analysis (GSEA)

Pathway enrichment analysis of the *cis-* and *trans*-eQTMs was performed using the gprofiler2 tool^87^. We obtained the observed overlap and the total term sizes for different pathways [GO:BP, KEGG, Reactome, Wiki pathways] to estimate the FE and the hypergeometric P value of enrichment for each pathway terms. Pathways were considered enriched after Bonferroni correction for multiple testing.

### Whole genome sequencing (WGS)

Whole genome sequencing (WGS) was carried out in the SG10K_Health participants using Illumina Hiseq X platform, as previously described^73^. Quality control (QC) and imputation of unmeasured genotypes were carried out, as described in Yew *et al*^88^. In brief, to address missingness created by the 15X sequencing, the TopMed Imputation Server was used to impute autosomal SNPs to the TopMed (Version R2) reference panel using the EAGLE2+Minimac4 pre-phasing and imputation pipeline for n=9,766 individuals in SG10K_Health. Approximately 285 million autosomal SNPs were available following imputation. Post-imputation quality control excluded imputed SNPs with MAF <0.001 in at least one of the three main ancestral groups (Chinese, Indians, and Malays), as well SNPs with imputation INFO score <0.30 and RUTH Hardy-Weinberg Equilibrium test^89^ P <10^-3^. A total of 7,857,631 imputed autosomal SNPs formed the final dataset, with 7,150,557 SNPs remaining with a MAF filter of 0.01.

### Identification of SNPs influencing the Sentinel CpGs

We carried out genome-wide analyses of the Sentinel CpGs to identify mQTL SNPs. Genetic association testing was done in 7,429 individuals from the five adult cohorts within SG10K_Health for which both DNA methylation and imputed genotype data was available. In brief, the methylation array data were quality controlled within each of the cohorts, adopting the identical approaches to those used in our epigenome-wide association analyses. Mixed Linear Model (MLM) based GWAS was performed to identify the genetic variants associated with the sentinel CpGs. The analysis was performed using the MLMA method implemented in GCTA tool^90^ while controlling for age, sex, ethnicity, proportion of six white blood cell sub-populations estimated by the Houseman method^76^, and the control probe PCs.

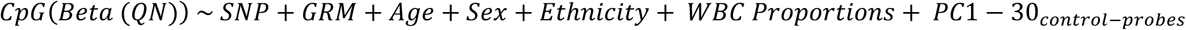

The *cis*-mQTL for each sentinel CpG was determined based on the strongest associated SNP significant at P< 2.7x10^-8^ (i.e. P<0.05 after Bonferroni correction for the number of *cis*- mQTLs tested). The *trans*-mQTLs were SNPs (at least beyond 10MB remote) from the CpG site and associated at P<5x10^-8^. We noted that multiple SNPs were associated with more than one CPG locus. To explore these pleotropic effects, we therefore quantified the number of CpGs associated with each mQTL SNP. We selected the Sentinel mQTL SNPs as being those with the highest number of associated sentinel CpGs and the lowest p-value for association with any sentinel CpG. We performed clumping of the *trans-*mQTLs into genomic loci based on 1MB intervals. This generated 242 independent Sentinel *trans*- mQTLs, each with at least one CpG associated.

The Variance explained and the heritability of the mQTLs for each CpG was calculated using the Genomic-relatedness-based Restricted Maximum-Likelihood (GREML) analysis^91^ implemented in the GCTA tool^90^. We calculated the variance explained by the top *cis*-mQTL, all cis-SNPs, and genome wide SNPs adjusting for the same covariates as used in the GWAS analysis. The R^2^ for each set was taken as the genetic variance explained by the SNPs (Vg), and the Heritability estimate was calculated as the ratio of genetic variance (Vg) to the total Phenotypic variance after adjusting for the covariates (Vp).

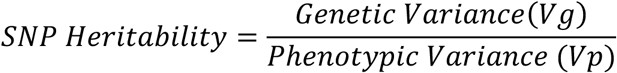

### Functional enrichment of cis- and trans-mQTLs

To examine the functional relevance of the mQTL SNP pathways to T2D, we tested whether the mQTLs were enriched for association with 16 key cardio-metabolic traits in published GWA studies. We determined the number of *cis-* and *trans*-mQTLs associated with the respective phenotypes at two thresholds (nominal: P<0.05; suggestive: P<1x10^-5^). To determine whether Sentinel mQTLs are enriched for phenotypic association compared to expectations under the null hypothesis, we determined background counts of phenotypic associations amongst 1000 random matched sets of SNPs (MAF and LD based matching). FE for each trait was calculated as the ratio of proportion of sentinel mQTLs associated to mean proportion of random SNPs associated and the P value was obtained from the distribution in the random sets.

We evaluated the (Observed) enrichment of the *trans*-mQTLs SNPs for association with multiple Sentinel CpGs, in comparison to expectations under the null hypothesis (Background), by hypergeometric testing. To determine Background expectations, we identified the number of Sentinel mQTLs, and the total number of CpGs associated with each mQTL, on a genome-wide basis across all quantified MethylEPIC array probes (n=837,222 CpGs). Fold enrichment (FE) for each mQTL was calculated as the ratio of proportion of Sentinel CpGs associated with an mQTL compared to the proportion of Background CpGs associated with an mQTL. The P value for enrichment was determined using the hypergeometric test, with Bonferroni correction applied for multiple testing (i.e. P<2x10^-4^, correction for N=242 *trans*-mQTLs tested).

### SMR and Colocalization analyses

We applied techniques to evaluate the potential causal relations of DNA methylation at the sentinel CpGs with T2D and other related metabolic phenotypes, as well as methylation with gene expression^32^. Cis- and trans-acting genetic variants influencing methylation were selected as described above (see GWAS of Sentinel CpGs in Methods). SNP-phenotype associations were from published GWAS studies (as annotated in the respective table), while SNP-gene expression (eQTLs) were from eQTLgen^34^. For loci whereby SMR estimates suggest a potential causal relationship for methylation at the sentinel CpGs on phenotype or gene expression at P<1.9x10-4 for cis-acting genetic variant (i.e. P<0.05 after Bonferroni correction for 264 CpGs) or at P<6.6x10-5 for trans-acting genetic variant (i.e. P<0.05 after Bonferroni correction for 759 SNP-CpG pairs), these were followed up with colocalization analysis (coloc v5.2.3)^33^ to assess if sufficient evidence exists for a shared genetic basis. For SMR , we performed single variant analyses using the Wald’s ratio-based MR calculation to infer causal relationships. For colocalization analysis, we assumed a single causal variant per locus with the default priors (P_1_ < 1x10^-4^; P_2_ < 1x10^-4^ and P_12_ < 1x10^-5^). As the array only covers 2-3% of all CpG sites in the epigenome, the biologically relevant CpGs site colocalises with the T2D might not be represented on the array. We therefore inferred potential colocalization based on either posterior probability PP3 (Shared loci, but different causal variant) or PP4 (Shared loci with same causal variant) greater than 0.6 as colocalised.

### Organisation of trans-mQTLs into potential networks

We clustered *trans*-mQTL SNPs based on their shared CpG sites, We started by creating a similarity matrix by calculating the ratio of shared CpGs compared to the total number of unique associated CpGs per SNP pair. We then computed a distance matrix representing the dissimilarity between each pair of SNPs (1 – similarity matrix). Hierarchical clustering was performed on the distance matrix using the average linkage method. Based on the silhouette width plot and the dendrogram visualization, we divided the 33 sentinel trans-mQTLs SNPs into clusters.

### Association of rs174598 (GàA) with plasma metabolite levels

The generation and QC of the plasma metabolite measurements has been described elsewhere^17,92^. Briefly, untargeted mass spectrometry based metabolomic profiling was done using the Metabolon HD4 panel. QC was performed to remove second visit samples, outliers and samples with greater than 25% missingness. Association of SNP rs174598 with the fatty acid metabolites were obtained from the trans-ancestry GWAS meta-analyses as done in the HELIOS study^17^. The individual GWAS for each ancestry was performed using REGENIE^93^ and the meta-analysis was performed using METAL^77^ with a fixed effect model controlling for genomic inflation across each dataset.

### SRE1 Binding motif scanning in the metabolic gene cluster

To identify whether the cis-eQTLs and the CpG-Genes in the metabolic gene cluster have a *SREBF1* binding site in their promoter region, we extracted the DNA sequence up to 3000bp upstream of each gene. We obtained the position weighted matrix (PWM) for the three SRE1 binding consensus sequences from the JASPAR database^94^ and used them alongside the sequences to find matching sequences across the different genes using the Find Individual Motif Occurrences (FIMO) scanning tool^95^ implemented in the MEME suite of motif based sequence analysis tools (https://meme-suite.org/)^96^. FIMO calculates the score of a motif match using the PWM, which represents the log-likelihood of observing a given base at each position of the motif and estimates a p-value representing the probability of observing a score at least as large as observed under the null hypothesis (random match with the background distribution). For all genes with at least one strong SRE1 consensus sequence, we determined the association with *SREBF1* expression adjusting for age, sex, ethnicity, RIN and the top six PEER factors.

### Whole genome bisulfite sequencing

We completed whole-genome bisulfite sequencing (WGBS) in 500 individuals (250 people with incident T2D and 250 controls) from amongst the LOLIPOP cohort. Cases were selected at random from amongst the available samples with incident T2D. Controls were matched to cases for age, gender, ethnic group and duration of follow-up. All participants were free from T2D at baseline. Aliquots of whole blood collected at enrolment, and stored at -80C, were used for extraction of genomic DNA. Two hundred nanograms of genomic DNA was used for bisulfite conversion with EZ DNA Methylation Gold Kit (Zymo Research), followed by library construction with the Accel-NGS® Methyl-Seq DNA Library Kit (Swift Biosciences), according to manufacturers’ protocols as previously described^41^. Each library was spiked in with 5% PhiX to compensate for the reduced sequence diversity in bisulfite converted libraries, with sequencing performed on an Illumina® NovaSeq 6000 platform to generate 2x150bp paired-end libraries. Data quality analysis and processing were performed as outlined in Zhou *et al*^41^. Reads are required to have a mapping quality (MAPQ) score >=5. Reads and generation of Picard metrics were performed with samtools v0.1.19. DNA methylated sites were identified, extracted and counted by bismark_methylation_extractor. The ENCODE blacklist was used to remove potentially dubious sites^97^, and sites with 1000 Genome South Asian (SAS) MAF of > 0.01 were removed using bedtools v2.30.0^98^ (**Extended Data Figure 8**). Read depth of coverage and insert size were analysed by Picard tools v2.25.2^99^ Effective depth was calculated after removing read duplicates, and counting overlapping bases only once. Only CpG sites with depth of coverage >5x were considered for further analysis. All data analyses were conducted by custom-made bash and R scripts (R version >= 3.4.4).

### Targeted methylation sequencing

We carried out targeted methylation for 2,000 samples from the iHealth-T2D cohort, comprising 988 incident T2D cases and 1,012 controls, using a custom library (TWIST Biosciences). Probes were designed, optimised, and synthesised using TWIST’s Custom Panel Design and Oligo synthesis solutions to target the 420 T2D associated genomic loci. We also included the 450 Houseman probes in the array design, to enable estimation of WBC cell type proportions. The total target region for methylation sequencing was 6.61Mb (**Supplementary Table 24**).

Sample processing and library preparation followed the TWIST Bioscience end-to-end targeted methylation sequencing workflow, as described in the product literature. For each sample, 200ng of genomic DNA underwent fragmentation to approximately 265 bp using the Bioruptor, end-repaired at 20°C and ligated with an EM-seq adapter, enzymatic methylation conversion (New England Biolabs® EMseqTM), and PCR amplification (9 cycles). Subsequently, 187.5ng of each enzymatically converted, amplified, and indexed library was hybridised to a custom methylation panel using the TWIST Bioscience Fast Hybridization and Wash Kit in accordance to the vendor’s protocols with the following modifications. Fast Hybridization was done for 2 hours at 60°C, followed by Fast Wash1 at 63°C and Fast Wash2 at 48°C. The final libraries were sequenced with the inclusion of negative and positive controls using the Illumina Novaseq platform, employing 150-bp paired-end reads.

Sequencing data was processed as previously described, with read trimming performed using Trim Galore (v0.6.4)^100^. Methylation levels at individual sites were called using MethylDackel (v0.5.3; –minDepth 10–maxVariantFrac 0.25–OT 0,0,0,138—OB 0,0,13,0). Three samples were removed from subsequent analyses due to low quality (one with mean target coverage <100x, two with poor concordance with respective EPIC data). Prior to association testing, 95% marker and sample call rate filtering were performed; and subsequently quantile normalised. This results in the remaining of 1,974 samples (978 cases and 996 controls).

### Methylation Risk Score (MRS) analyses

#### Array-based MRS

Array-based MRS were developed using data from the MEC study (Development set). Model performance was then evaluated in the iHealth-T2D samples (Test set) to provide independent evaluation. We built the model based using only the 102 sentinel CpGs associated with T2D at P<0.05 and spearman correlation of R <0.8 in the MEC study (Development set). These were entered into the LASSO model for variable selection^101^. A 10-fold cross validation with “lambda.1se” criterion was performed to determine the most optimal lambda (λ) where the value of λ represented the most regularised model in which the error was within one standard error of the minimum. The MRS was calculated in iHealth- T2D study (Test set) using the linear combination of z-scaled methylation values and the nonzero coefficients retained from the LASSO regression in MEC.

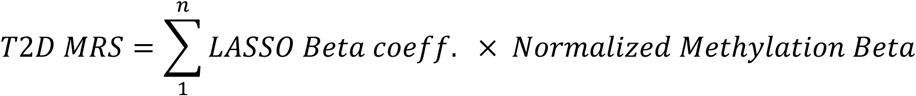

### Gene-based MRS based on targeted sequencing

We carried out complete gene methylation sequencing at the *ABCG1* and *SREBF1* loci, as described. We found methylation at 21 CpG sites to be associated with T2D at the *ABCG1* locus, and 5 at the *SREBF1* locus The P value thresholds for selection were P<2.6x10^-6^ for *ABCG1* and P<3.1x10^-6^ for *SREBF1*, representing P<0.05 after Bonferroni correction for the 19,217 and 16,283 CpGs sequenced at the *ABCG1* and *SREBF1* locus respectively. We entered these CpGs into LASSO modelling to create gene-based MRS. We split the fine-mapping dataset into 70:30 for Development and Testing. Within the Development set, we further undertook a 10-fold cross-validation approach to obtain the optimum model, which was then tested in the remaining 30% of the dataset (Test set). This resulted in 17 and five CpGs selected by LASSO for inclusion in our gene-based MRS model, at *ABCG1* and *SREBF1* respectively.

### Sequenced-based MRS

For our sequence-based MRS, we included 165 CpGs within the primary target region that were associated with T2D at P<3.7x10^-6^, representing P<0.05 after Bonferroni correction for the 13,338 CpGs sequenced within primary target region, of which 60 were selected by LASSO. Similar to the array- and gene-based MRS, we split the dataset 70:30 for Development and Testing, with a 10-fold cross-validation approach within the Development set.

## Code and Data availability

All genomic positions are provided as GRCh38 co-ordinates. The analytic codes are available in the study <u>GitHub repository</u>. The complete association statistics for the EPIC array based CpG sites and the targeted sequencing based CpG sites with Type 2 Diabetes are also available [upon publication] via our github, along with summary statistics for the *cis*- mQTL, *trans*-mQTL, *cis*-eQTM and *trans*-eQTM analyses. Access to individual level phenotype and molecular data used in this manuscript can be obtained by contacting the respective cohort Data Access Committees.

## Data Availability

All genomic positions are provided as GRCh38 co-ordinates. The analytic codes are
available in the study GitHub repository. The complete association statistics for the EPIC
array based CpG sites and the targeted sequencing based CpG sites with Type 2 Diabetes
are also available [upon publication] via our github, along with summary statistics for the cismQTL,trans-mQTL, cis-eQTM and trans-eQTM analyses. Access to individual level
phenotype and molecular data used in this manuscript can be obtained by contacting the
respective cohort Data Access Committees.

## Acknowledgements and Funding

This research is supported by the Singapore Ministry of Health’s National Medical Research Council under its Singapore Translational Research (STaR) Investigator. The HELIOS study is supported by the Singapore Ministry of Health’s National Medical Research Council under its OF-LCG funding scheme (NMRC Project Ref. MOH-000271-00), STaR funding scheme (NMRC Project Ref. NMRC/STaR/0028/2017) and intramural funding from Nanyang Technological University, Lee Kong Chian School of Medicine and the National Healthcare Group. The Multi-Ethnic Cohort Phase 1 (MEC1) are supported by individual research and clinical scientist award schemes from the National Medical Research Council (NMRC) and the Biomedical Research Council (BMRC) of Singapore, and infrastructure funding from the Singapore Ministry of Health (Population Health Metrics and Analytics PHMA), National University of Singapore and National University Health System, Singapore. The LOLIPOP study was funded by the National Institute for Health Research (NIHR) (16/136/68) using UK aid from the UK Government to support global health research, and by Wellcome Trust (212945/Z/18/Z). The views expressed in this publication are those of the author(s) and not necessarily those of the NIHR or the UK Department of Health and Social Care. The SCHS was funded by the National Medical Research Council, Singapore [NMRC/CIRG/1354/2013] and National Institutes of Health, USA [RO1 CA144034 and UM1 CA182876]. We thank all participants, and members of the study team, for their research contributions.RNA sequencing was partially funded by i) Ministry of Education Academic Research Fund Tier 1 Grant (RS09/20), ii) A*Supplementary Table AR-NHMRC Joint Grant Call (A20PRb0138), iii) Start-Up Grant (awarded to Marie Loh [PI]) from Lee Kong Chian School of Medicine, Nanyang Technological University, Singapore and iv) Imperial - Nanyang Technological University Collaboration Fund (awarded to Marie Loh [PI]). The computational work for this study was partially performed on resources of the National Supercomputing Centre, Singapore (https://www.nscc.sg).

This study made use of the SG10K_Health dataset generated as part of the Singapore National Precision Medicine program funded by the Industry Alignment Fund (Pre-Positioning) (IAF-PP: H17/01/a0/007). The SG10K_Health dataset was created from data / samples collected in the following cohorts in Singapore: (1) The Health for Life in Singapore (HELIOS) study at the Lee Kong Chian School of Medicine, Nanyang Technological University, Singapore (supported by grants from a Strategic Initiative at Lee Kong Chian School of Medicine, the Singapore Ministry of Health (MOH) under its Singapore Translational Research Investigator Award (NMRC/STaR/0028/2017) and the IAF-PP: H18/01/a0/016); (2) The Growing up in Singapore Towards Healthy Outcomes (GUSTO) study, which is jointly hosted by the National University Hospital (NUH), KK Women’s and Children’s Hospital (KKH), the National University of Singapore (NUS) and the Singapore Institute for Clinical Sciences (SICS), Agency for Science Technology and Research (A*STAR) (supported by the Singapore National Research Foundation under its Translational and Clinical Research (TCR) Flagship Programme and administered by the Singapore Ministry of Health’s National Medical Research Council (NMRC), Singapore - NMRC/TCR/004-NUS/2008; NMRC/TCR/012-NUHS/2014. Additional funding is provided by SICS and IAF-PP H17/01/a0/005); (3) The Singapore Epidemiology of Eye Diseases (SEED) cohort at Singapore Eye Research Institute (SERI) (supported by NMRC/CIRG/1417/2015; NMRC/CIRG/1488/2018; NMRC/OFLCG/004/2018); (4) The Multi-Ethnic Cohort (MEC) cohort (supported by NMRC grant 0838/2004; BMRC grant 03/1/27/18/216; 05/1/21/19/425; 11/1/21/19/678, Ministry of Health, Singapore, National University of Singapore and National University Health System, Singapore); (5) The SingHealth Duke-NUS Institute of Precision Medicine (PRISM) cohort (supported by NMRC/CG/M006/2017_NHCS; NMRC/STaR/0011/2012, NMRC/STaR/ 0026/2015, Lee Foundation and Tanoto Foundation); (6) The TTSH Personalised Medicine Normal Controls (TTSH) cohort funded (supported by NMRC/CG12AUG17 and CGAug16M012). The views expressed are those of the author(s) are not necessarily those of the National Precision Medicine investigators, or institutional partners. We thank all investigators, staff members and study participants who made the National Precision Medicine Project possible.

---

**Extended Data Figure 1.**
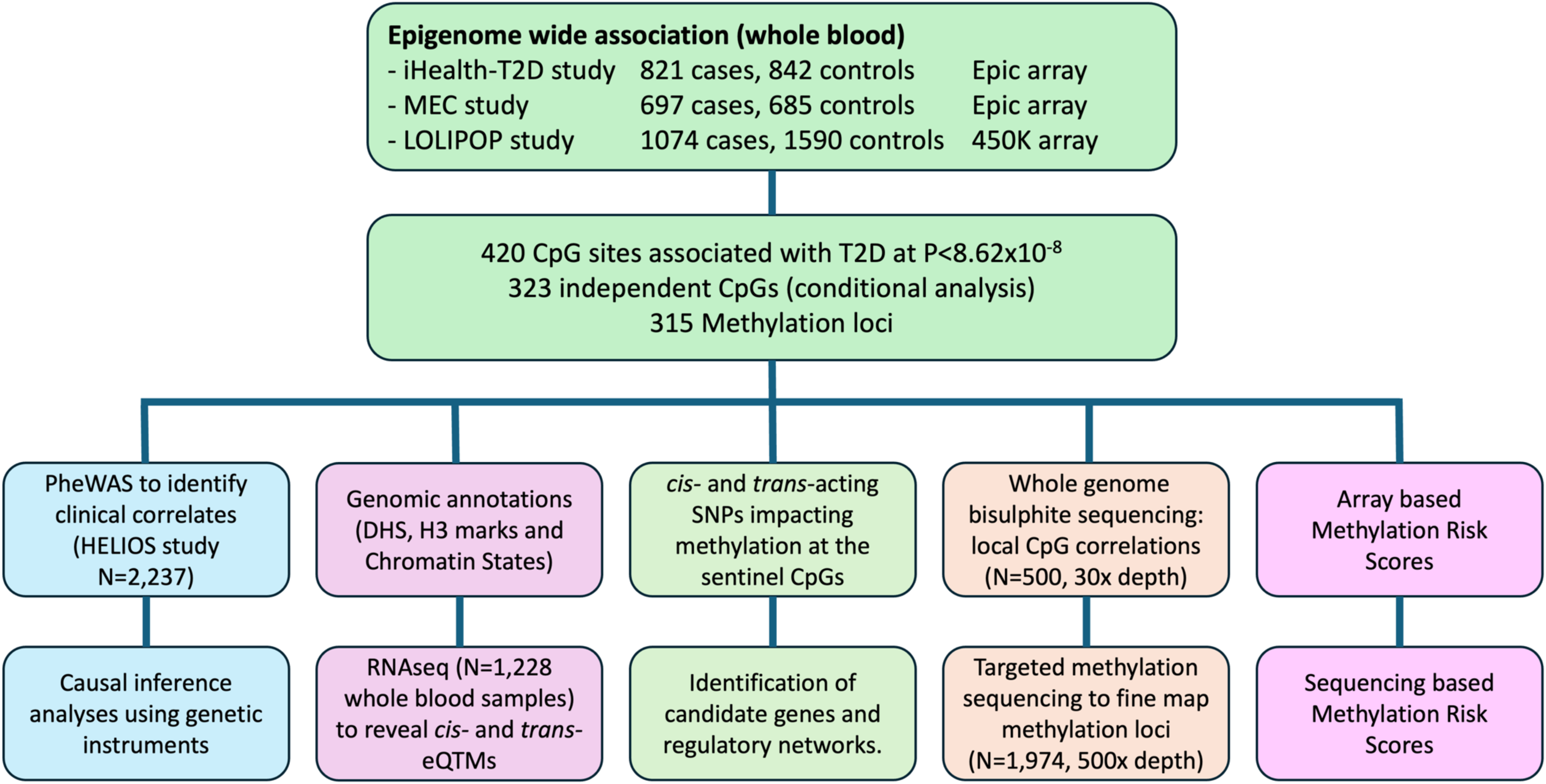
Overview of the study design.

**Extended Data Figure 2:**
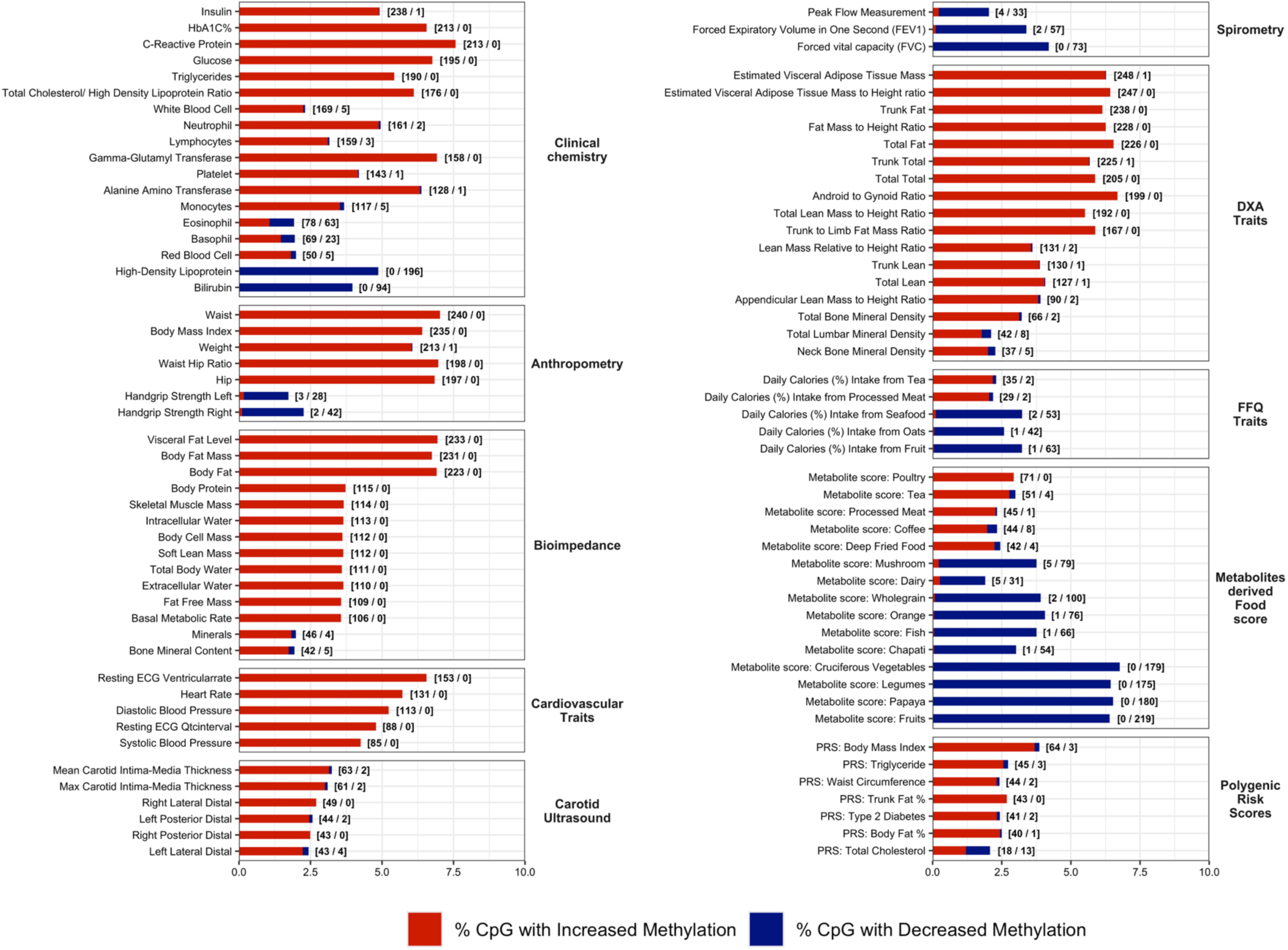
Phenome-wide association of the Sentinel CpGs with epidemiological exposures, amongst participants of the HELIOS study. The significant associations with permutation P-value <0.001 across the different categories are shown. The x axis represents the Fold enrichment and the y-axis are the individual traits split by their category. The colour represents the percentage of sentinel CpGs associated with an increased risk (red) and decreased risk (blue) for each trait. The numbers next to the plot show the count of CpGs associated with increased / decreased risk respectively.

**Extended Data Figure 3.**
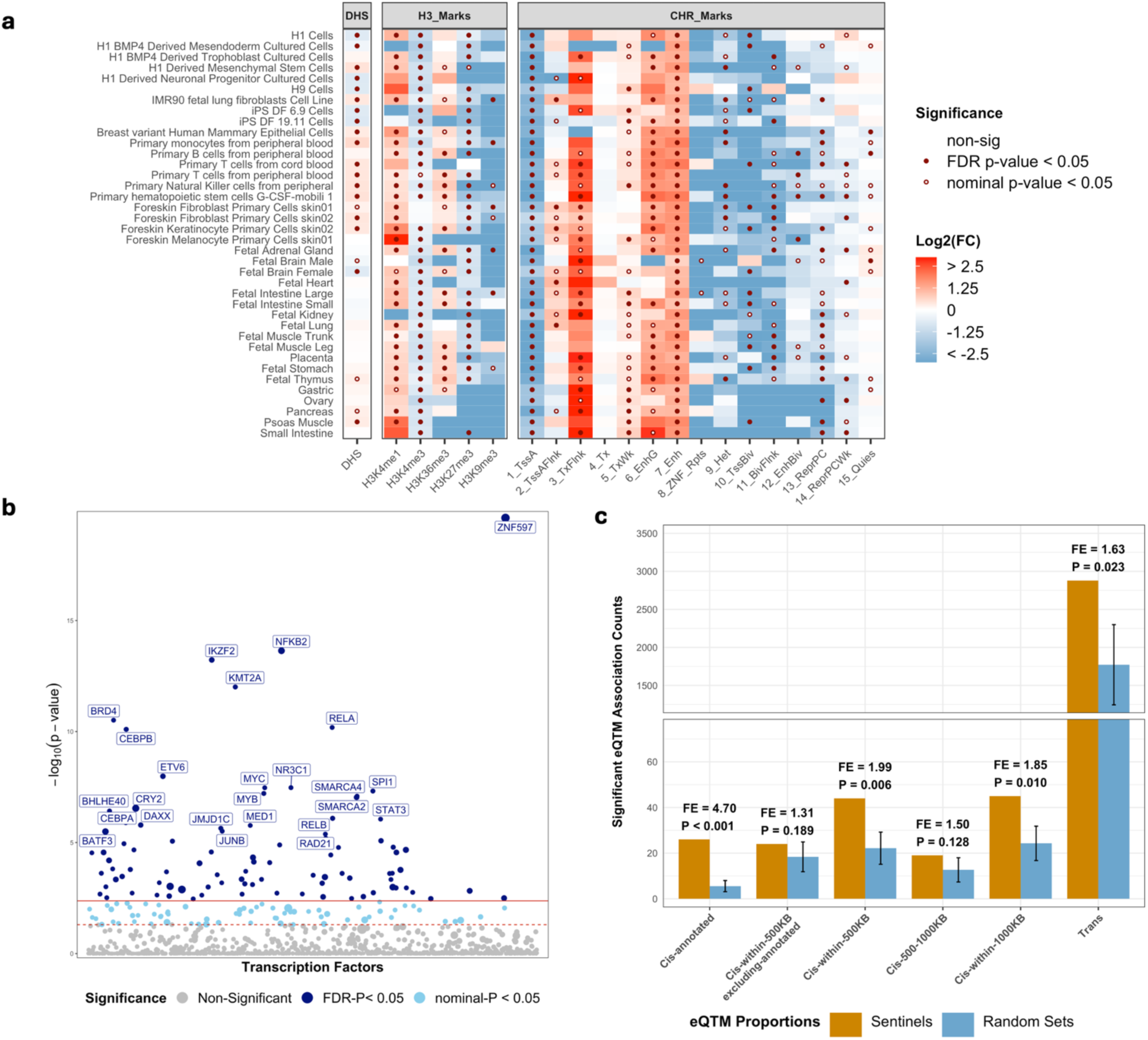
Functional Enrichment of Sentinel CpGs. **Panel a)** Functional annotation and enrichment of Sentinel CpGs across different cell types. Enrichment is shown as observed count vs expected background count across DNase 1 Hotspots (DHS); five Histone 3 marks and 15 Chromatin States. **b)** Enrichment of Sentinel CpGs across 1210 transcription factors (TFs) from the ReMAP database. The top 25 significantly enriched TFs are labelled. **c)** Enrichment of Sentinel CpG associated genes both in cis and trans. Cis-genes were annotated using 5 different threshold criteria to identify the extent of enrichment compared to the nearest genes vs alternate choices of gene sets within the 1MB region.

**Extended Data Figure 4.**
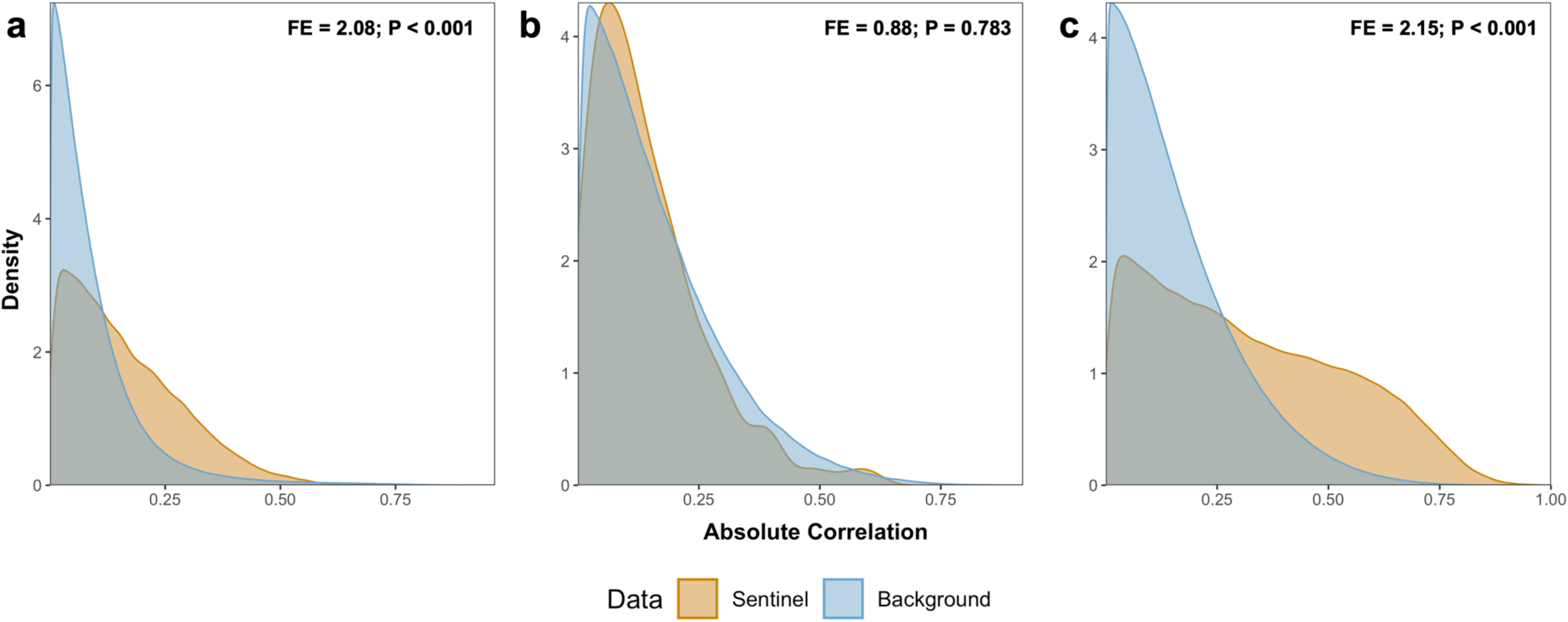
Covariation of Sentinel CpG and their associated eQTM Signatures. Pairwise absolute correlation between **a)** Sentinel CpGs; **b)** cis-eQTMs (nearest gene only); and **c)** trans-eQTM. Covariation in 1000 random Background sets is shown for comparison, and for probability estimation. The fold enrichment was calculated as the ratio of mean absolute correlation in the Sentinel CpG set compared to the mean in the background sets. P-value for enrichment was obtained using a two-sided t-test.

**Extended Data Figure 5.**
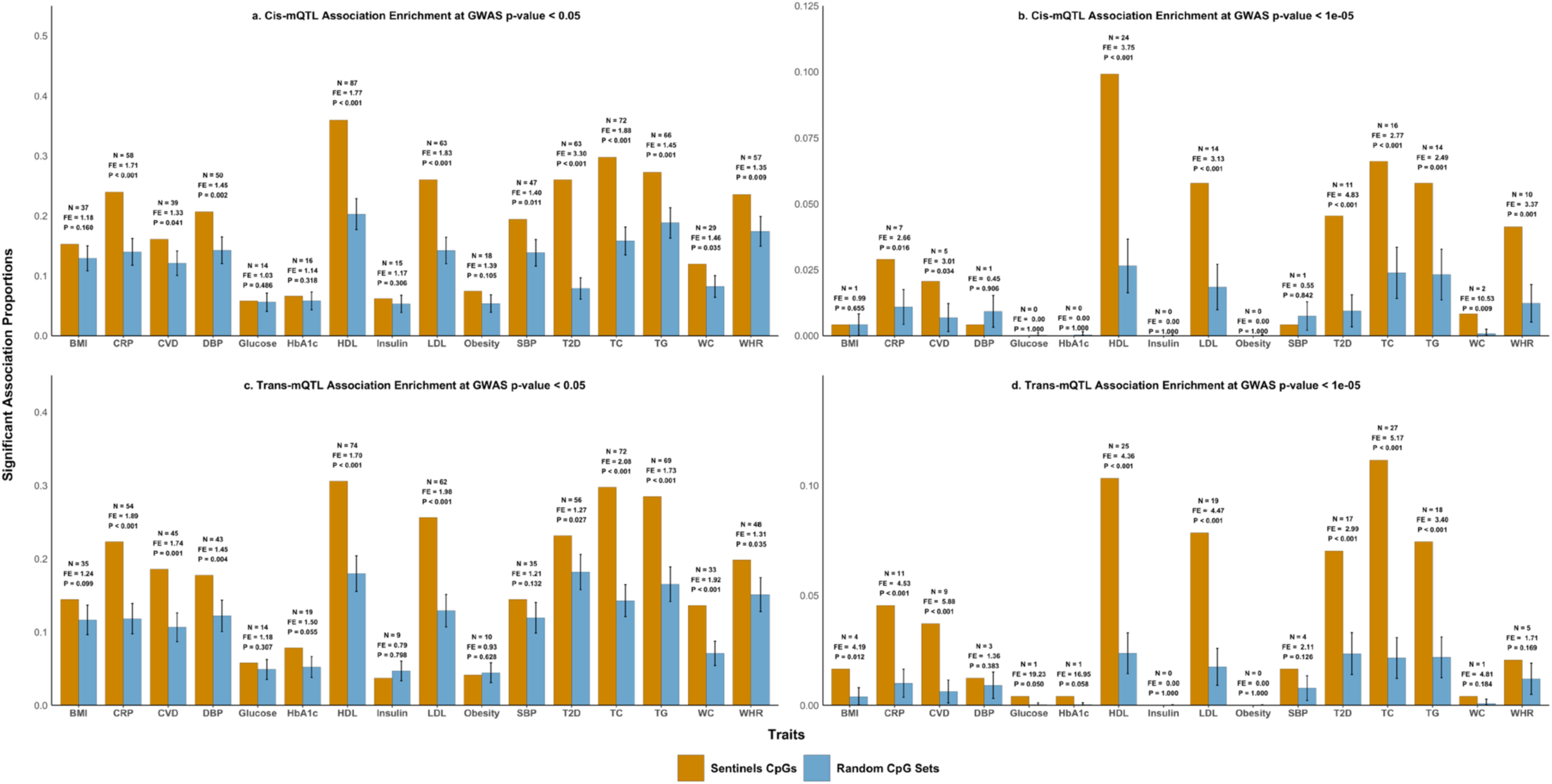
Enrichment analysis: *cis*- and *trans*-acting mQTL SNPs that influence Sentinel CpGs, are enriched for association with T2D and other related human metabolic traits. Proportion of Sentinel *cis*-acting mQTL SNPs associated with cardiometabolic traits compared to background, at a GWAS threshold of P<0.05 (**a**) or at P<1x10^-5^ (**b**). Proportion of Sentinel *trans-*acting mQTL SNPs associated with cardiometabolic traits compared to background, at a GWAS threshold of P<0.05 (**c**) or at P<10^-5^ (**d**).

**Extended Data Figure 6.**
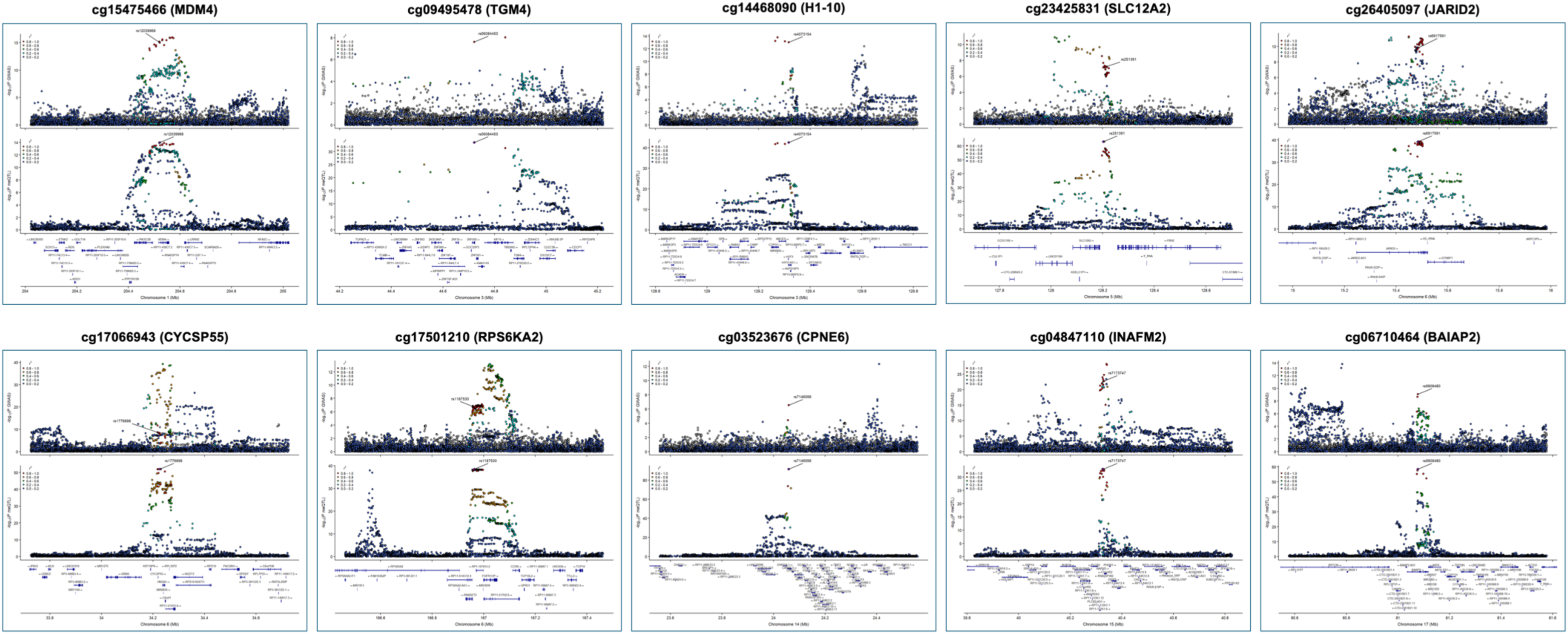
Cis-mQTL based colocalization analysis of Sentinel CpGs and Type 2 Diabetes. Regional plots of associations between the ten sentinel CpG loci that have a potential casual association with T2D risk using SMR and colocalization analyses. The lead *cis*-mQTL for each CpG site which is used as the instrument variable is labelled and the other SNPs are coloured based on their LD correlation with the lead SNP.

**Extended Data Figure 7.**
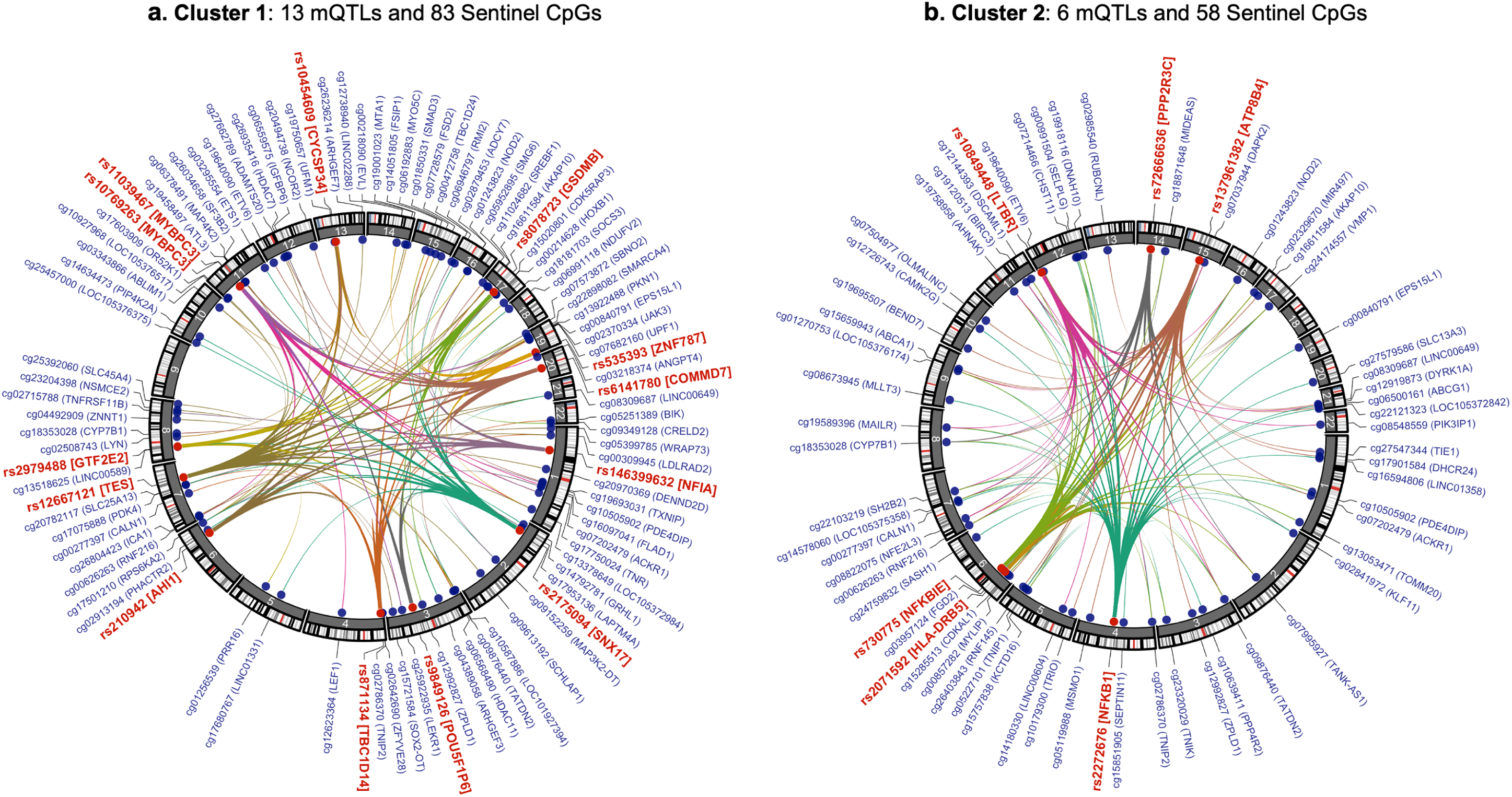
*Trans*-acting SNP-CPG clusters. Circos plot illustrating the two largest *trans-*mQTL clusters influencing methylation at Sentinel CpGs**. Panel a)** Cluster 1: 13 Sentinel *trans-*mQTL SNPs and 83 CpGs. **b)** Cluster 2: six *trans-*mQTL and 58 CpGs. The outer track provides the SNP rsIDs (red text) and CpG ID (blue text). SNPs are and CpGs are additionally annotated with the cis-eQTL most closely associated with the SNP, and gene closest to the CpG. Inner connections show the trans-acting associations between mQTL SNPs and respective CpGs. The connections are colour coded according to the respective *trans*-mQTL. The identified *cis-*eQTLs include *NKFB1, NFKBIA, NFKBIE, NF1A, COMMD7, IKZF3, MADD* and *MYBPC3*. Many of the genes at the linked *trans*-Sentinel CpG sites also encode recognised inflammatory mediators (eg *JAK3, MAP3K2, NOD2, SMAD3, TGFBR1* and *TNIP1*). However, the networks also link to CpG-genes that are components of metabolic pathways directly relevant to the pathophysiology of diabetes, including *CDKAL1, CPT1A, CYP7B1, PDK4, LDLRAD2, SREBF1, SH2B2, SOCS3, TANK* and *TXNIP*. These genes are reported to impact pancreatic beta cell function, insulin signalling and action, glucose sensing, metabolism of glucose, cholesterol and lipids, fatty acid beta oxidation, mitochondrial biology, thermogenesis, and adipogenesis, and are thus compelling candidate genes in the pathogenesis of diabetes.

**Extended Data Figure 8.**
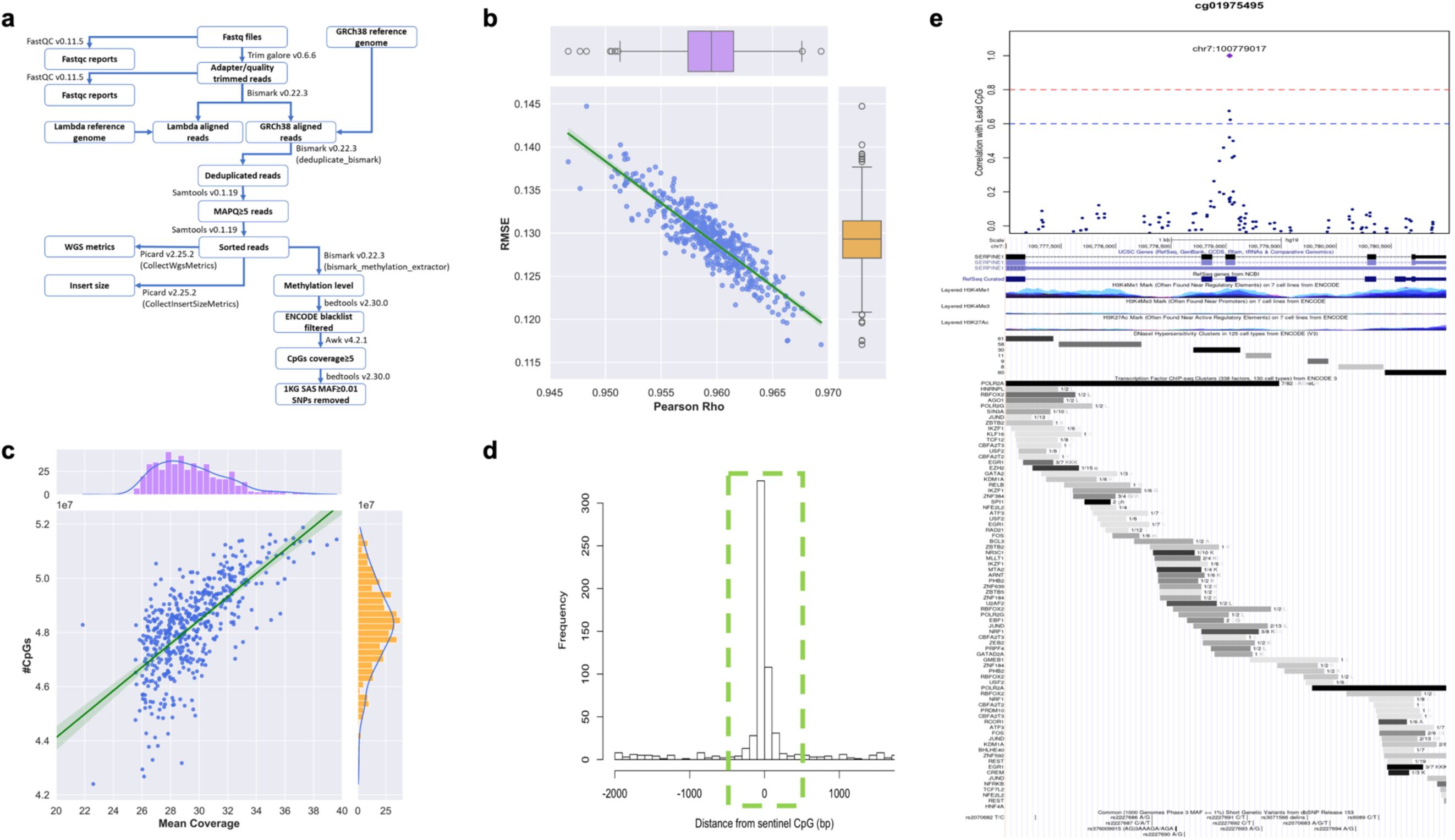
WGBS Pipeline for Data generation, curation and quality control. **Panel a)** analytics pipeline used to process the WGBS samples. **b)** Joint plot comparing the common methylation CpG signals between WGBS and its corresponding 450K array across 500 samples – average (S.D.) number of CpGs compared is 351,684.3 (± 22,035.5). The main diagram depicts the Pearson Rho and RMSE of the methylation beta value compared, while the top and right joint diagrams portray the boxplot distribution of the Pearson Rho and RMSE values respectively. **c)** Demonstrates the distribution of the mean coverage and number of CpGs across the 500 WGBS samples. The top and right joint diagrams provide the histogram distribution of the mean coverage and number of CpGs respectively. **d)** Histogram for the distribution of distance of most outlying CpG with |r|>0.2 within +/-2kb from corresponding discovery CpGs **e)** Example of local correlation structure with discovery CpG (array) from WGBS data, overlaid with UCSC and RefSeq genes and regulatory features (histone marks, DHS clusters, TF from ChIP-seq)

**Extended Data Figure 9.**
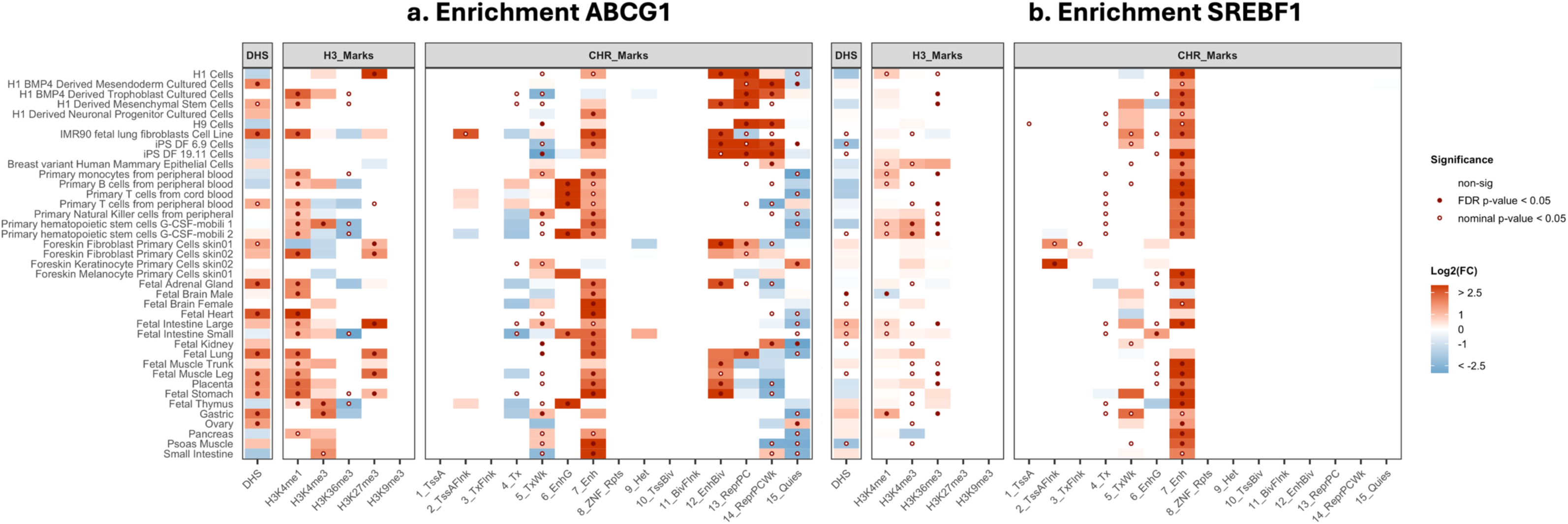
Functional Enrichment of the Fine-mapping loci. Functional annotation and enrichment of T2D associated methylation sites in the *ABCG1* Fine-mapped region **(a)** and the SREBF1 fine-mapped region **(b)** , compared to all the CpGs in each of the two regions respectively. Enrichment is shown as the ratio observed count vs expected background count across DNase 1 Hotspots (DHS); five Histone 3 marks and 15 Chromatin States in different cell types.

**Extended Data Figure 10.**
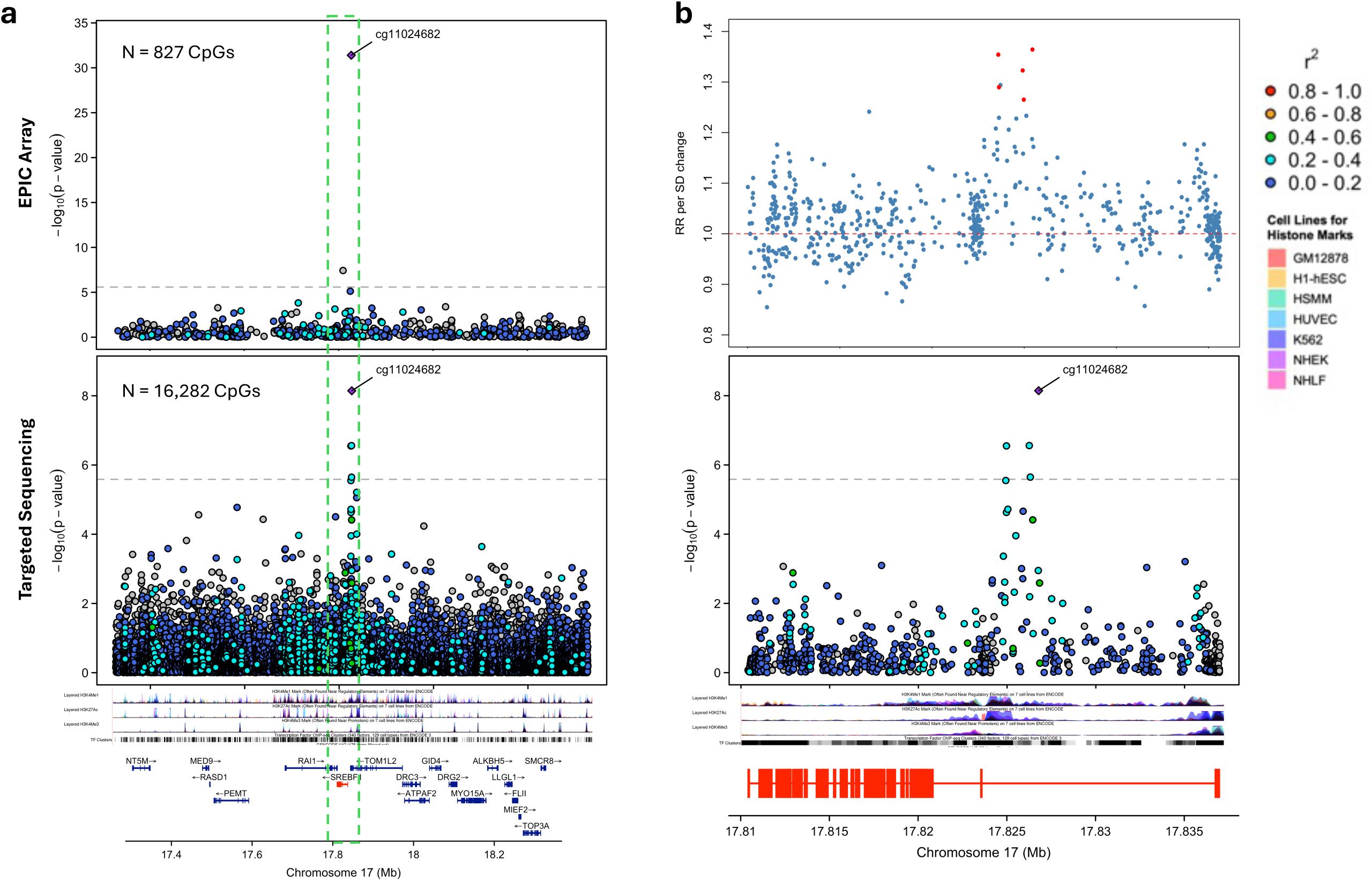
Fine-mapping at *SREBF1* locus. **Panel a)** Top and bottom panel shows the -log10(p) value of association with T2D for CpG sites captured by the EPIC array and TWIST targeted sequencing respectively within a 1Mb region around the sentinel CpG (cg11024682). **b)** Zoomed in regional plot of *SREBF1* genic region, as indicated green rectangle in. Top panel shows the relative risk (RR) for T2D per standard deviation (SD) change in methylation level, whilst the lower panel indicates the -log10(p) value of association with T2D for CpG sites within the *SREBF1* genic region. The correlation with the index CpG is highlighted using the different colors. Information about the regulatory regions was obtained from UCSC genome browser for seven cell lines and are highlighted and labelled in the legend. (GM12878: Lymphoblastoid cells; H1-hESC: H1 human embryonic stem cell line; HSMM: Human skeletal muscle myoblasts; HUVEC: Human umbilical vein endothelial cells; K562: human chronic myelogenous leukemia (CML) cell line; NHEK: Normal Human Epidermal Keratinocytes; NHLF: Normal human Lung Fibroblasts)

## References

1. NCD Risk Factor Collaboration (NCD-RisC). Worldwide trends in diabetes prevalence and treatment from 1990 to 2022: a pooled analysis of 1108 population-representative studies with 141 million participants. Lancet Lond. Engl. 404, 2077–2093 (2024).

2. Sun, H. et al. IDF Diabetes Atlas: Global, regional and country-level diabetes prevalence estimates for 2021 and projections for 2045. Diabetes Res. Clin. Pract. 183, 109119 (2022).

3. Anjana, R. M. et al. Metabolic non-communicable disease health report of India: the ICMR-INDIAB national cross-sectional study (ICMR-INDIAB-17). Lancet Diabetes Endocrinol. 11, 474–489 (2023).

4. Chambers, J. C. et al. Epigenome-wide association of DNA methylation markers in peripheral blood from Indian Asians and Europeans with incident type 2 diabetes: a nested case-control study. Lancet Diabetes Endocrinol. 3, 526–534 (2015).

5. Cheng, Y. J. et al. Prevalence of Diabetes by Race and Ethnicity in the United States, 2011-2016. JAMA **322**, 2389–2398 (2019).

6. Sattar, N. & Gill, J. M. R. Type 2 diabetes in migrant south Asians: mechanisms, mitigation, and management. Lancet Diabetes Endocrinol. 3, 1004–1016 (2015).

7. Ke, C., Narayan, K. M. V., Chan, J. C. N., Jha, P. & Shah, B. R. Pathophysiology, phenotypes and management of type 2 diabetes mellitus in Indian and Chinese populations. Nat. Rev. Endocrinol. 18, 413–432 (2022).

8. Cefalu, W. T. et al. A global initiative to deliver precision health in diabetes. Nat. Med. 30, 1819–1822 (2024).

9. Yang, H. et al. The association between sodium-glucose cotransporter 2 inhibitors and contrast-associated acute kidney injury in patients with type 2 diabetes undergoing angiography: a propensity-matched study. Eur. J. Med. Res. 29, 621 (2024).

10. Florath, I. et al. Type 2 diabetes and leucocyte DNA methylation: an epigenome-wide association study in over 1,500 older adults. Diabetologia 59, 130–138 (2016).

11. Cardona, A. et al. Epigenome-Wide Association Study of Incident Type 2 Diabetes in a British Population: EPIC-Norfolk Study. Diabetes 68, 2315–2326 (2019).

12. Albao, D. S. et al. Methylation changes in the peripheral blood of Filipinos with type 2 diabetes suggest spurious transcription initiation at TXNIP. Hum. Mol. Genet. 28, 4208– 4218 (2019).

13. Juvinao-Quintero, D. L. et al. DNA methylation of blood cells is associated with prevalent type 2 diabetes in a meta-analysis of four European cohorts. Clin. Epigenetics 13, 40 (2021).

14. Fraszczyk, E. et al. Epigenome-wide association study of incident type 2 diabetes: a meta-analysis of five prospective European cohorts. Diabetologia 65, 763–776 (2022).

15. Tan, K. H. X. et al. Cohort Profile: The Singapore Multi-Ethnic Cohort (MEC) study. Int. J. Epidemiol. 47, 699–699j (2018).

16. Edgington, E., Edgington, E. & Onghena, P. Randomization Tests. (Chapman and Hall/CRC, 2007). doi:10.1201/9781420011814.

17. Wang, X. et al. The Health for Life in Singapore (HELIOS) Study: delivering Precision Medicine research for Asian populations. Preprint at 10.1101/2024.05.14.24307259 (2024).

18. Hoffmann, A. et al. A polyphenol-rich green Mediterranean diet enhances epigenetic regulatory potential: the DIRECT PLUS randomized controlled trial. Metabolism. 145, 155594 (2023).

19. Guirette, M. et al. Genome-Wide Interaction Analysis With DASH Diet Score Identified Novel Loci for Systolic Blood Pressure. Hypertens. Dallas Tex 1979 81, 552–560 (2024).

20. Kresovich, J. K., Park, Y.-M. M., Keller, J. A., Sandler, D. P. & Taylor, J. A. Healthy eating patterns and epigenetic measures of biological age. Am. J. Clin. Nutr. 115, 171– 179 (2022).

21. Low, D. Y. et al. Metabolic variation reflects dietary intake in a multi-ethnic Asian population. Preprint at 10.1101/2023.12.04.23299350 (2023).

22. Gaziano, J. M. et al. Million Veteran Program: A mega-biobank to study genetic influences on health and disease. J. Clin. Epidemiol. 70, 214–223 (2016).

23. Pulit, S. L. et al. Meta-analysis of genome-wide association studies for body fat distribution in 694 649 individuals of European ancestry. Hum. Mol. Genet. 28, 166– 174 (2019).

24. Chen, J. et al. The trans-ancestral genomic architecture of glycemic traits. Nat. Genet. 53, 840–860 (2021).

25. Jiang, L., Zheng, Z., Fang, H. & Yang, J. A generalized linear mixed model association tool for biobank-scale data. Nat. Genet. 53, 1616–1621 (2021).

26. Said, S. et al. Genetic analysis of over half a million people characterises C-reactive protein loci. Nat. Commun. 13, 2198 (2022).

27. Aragam, K. G. et al. Discovery and systematic characterization of risk variants and genes for coronary artery disease in over a million participants. Nat. Genet. 54, 1803– 1815 (2022).

28. Graham, S. E. et al. The power of genetic diversity in genome-wide association studies of lipids. Nature 600, 675–679 (2021).

29. Chen, C.-Y. et al. Analysis across Taiwan Biobank, Biobank Japan, and UK Biobank identifies hundreds of novel loci for 36 quantitative traits. Cell Genomics **3**, 100436 (2023).

30. Yang, M.-L. et al. Sex-specific genetic architecture of blood pressure. Nat. Med. 30, 818–828 (2024).

31. Suzuki, K. et al. Genetic drivers of heterogeneity in type 2 diabetes pathophysiology. Nature 627, 347–357 (2024).

32. Zhu, Z. et al. Integration of summary data from GWAS and eQTL studies predicts complex trait gene targets. Nat. Genet. 48, 481–487 (2016).

33. Giambartolomei, C. et al. Bayesian test for colocalisation between pairs of genetic association studies using summary statistics. PLoS Genet. 10, e1004383 (2014).

34. Võsa, U. et al. Large-scale cis- and trans-eQTL analyses identify thousands of genetic loci and polygenic scores that regulate blood gene expression. Nat. Genet. 53, 1300– 1310 (2021).

35. Horton, J. D., Goldstein, J. L. & Brown, M. S. SREBPs: activators of the complete program of cholesterol and fatty acid synthesis in the liver. J. Clin. Invest. 109, 1125– 1131 (2002).

36. Leonardi, R., Rehg, J. E., Rock, C. O. & Jackowski, S. Pantothenate kinase 1 is required to support the metabolic transition from the fed to the fasted state. PloS One 5, e11107 (2010).

37. Benhammou, J. N. et al. Novel Lipid Long Intervening Noncoding RNA, Oligodendrocyte Maturation-Associated Long Intergenic Noncoding RNA, Regulates the Liver Steatosis Gene Stearoyl-Coenzyme A Desaturase As an Enhancer RNA. Hepatol. Commun. 3, 1356–1372 (2019).

38. Rigotti, A. et al. A targeted mutation in the murine gene encoding the high density lipoprotein (HDL) receptor scavenger receptor class B type I reveals its key role in HDL metabolism. Proc. Natl. Acad. Sci. U. S. A. 94, 12610–12615 (1997).

39. Zerenturk, E. J., Sharpe, L. J., Ikonen, E. & Brown, A. J. Desmosterol and DHCR24: unexpected new directions for a terminal step in cholesterol synthesis. Prog. Lipid Res. 52, 666–680 (2013).

40. Pidsley, R. et al. Critical evaluation of the Illumina MethylationEPIC BeadChip microarray for whole-genome DNA methylation profiling. Genome Biol. 17, 208 (2016).

41. Zhou, L. et al. Systematic evaluation of library preparation methods and sequencing platforms for high-throughput whole genome bisulfite sequencing. Sci. Rep. 9, 10383 (2019).

42. Giroud, M. et al. HAND2 is a novel obesity-linked adipogenic transcription factor regulated by glucocorticoid signalling. Diabetologia 64, 1850–1865 (2021).

43. Wahl, S. et al. Epigenome-wide association study of body mass index, and the adverse outcomes of adiposity. Nature 541, 81–86 (2017).

44. Smith, Z. D., Hetzel, S. & Meissner, A. DNA methylation in mammalian development and disease. Nat. Rev. Genet. 26, 7–30 (2025).

45. Daily, J. W. & Park, S. Association of Plant-Based and High-Protein Diets with a Lower Obesity Risk Defined by Fat Mass in Middle-Aged and Elderly Persons with a High Genetic Risk of Obesity. Nutrients 15, 1063 (2023).

46. Imamura, M. et al. Genome-wide association studies in the Japanese population identify seven novel loci for type 2 diabetes. Nat. Commun. 7, 10531 (2016).

47. Wang, X. et al. Interactive associations of the INAFM2 rs67839313 variant and egg consumption with type 2 diabetes mellitus and fasting blood glucose in a Chinese population: A family-based study. Gene 770, 145357 (2021).

48. Zhang, Y., Zeng, S. X., Hao, Q. & Lu, H. Monitoring p53 by MDM2 and MDMX is required for endocrine pancreas development and function in a spatio-temporal manner. Dev. Biol. 423, 34–45 (2017).

49. Cervantes, S. et al. Late-stage differentiation of embryonic pancreatic β-cells requires Jarid2. Sci. Rep. 7, 11643 (2017).

50. Abdelgawad, R. et al. Loss of Slc12a2 specifically in pancreatic β-cells drives metabolic syndrome in mice. PloS One 17, e0279560 (2022).

51. Rohm, T. V., Meier, D. T., Olefsky, J. M. & Donath, M. Y. Inflammation in obesity, diabetes, and related disorders. Immunity 55, 31–55 (2022).

52. Kakiyama, G. et al. Insulin resistance dysregulates CYP7B1 leading to oxysterol accumulation: a pathway for NAFL to NASH transition. J. Lipid Res. 61, 1629–1644 (2020).

53. Pedroso, J. A. B., Ramos-Lobo, A. M. & Donato, J. SOCS3 as a future target to treat metabolic disorders. Horm. Athens Greece 18, 127–136 (2019).

54. Ghosh, C. et al. Involvement of Cdkal1 in the etiology of type 2 diabetes mellitus and microvascular diabetic complications: a review. J. Diabetes Metab. Disord. 21, 991– 1001 (2022).

55. Saltiel, A. R. Insulin signaling in health and disease. J. Clin. Invest. 131, e142241, 142241 (2021).

56. Park, B.-Y. et al. PDK4 Deficiency Suppresses Hepatic Glucagon Signaling by Decreasing cAMP Levels. Diabetes 67, 2054–2068 (2018).

57. Bodur, C. et al. TBK1-mTOR Signaling Attenuates Obesity-Linked Hyperglycemia and Insulin Resistance. Diabetes 71, 2297–2312 (2022).

58. Wang, X., Shen, X., Yan, Y. & Li, H. Pyruvate dehydrogenase kinases (PDKs): an overview toward clinical applications. Biosci. Rep. 41, BSR20204402 (2021).

59. Thielen, L. A. et al. Identification of an Anti-diabetic, Orally Available Small Molecule that Regulates TXNIP Expression and Glucagon Action. Cell Metab. 32, 353–365.e8 (2020).

60. Hedman, Å. K. et al. Epigenetic Patterns in Blood Associated With Lipid Traits Predict Incident Coronary Heart Disease Events and Are Enriched for Results From Genome-Wide Association Studies. Circ. Cardiovasc. Genet. 10, e001487 (2017).

61. Hillary, R. F. et al. Blood-based epigenome-wide analyses of chronic low-grade inflammation across diverse population cohorts. Cell Genomics 4, 100544 (2024).

62. Wielscher, M. et al. DNA methylation signature of chronic low-grade inflammation and its role in cardio-respiratory diseases. Nat. Commun. 13, 2408 (2022).

63. Gomez-Alonso, M. D. C. et al. DNA methylation and lipid metabolism: an EWAS of 226 metabolic measures. Clin. Epigenetics 13, 7 (2021).

64. Kennedy, M. A. et al. ABCG1 has a critical role in mediating cholesterol efflux to HDL and preventing cellular lipid accumulation. Cell Metab. 1, 121–131 (2005).

65. Yvan-Charvet, L. et al. Combined deficiency of ABCA1 and ABCG1 promotes foam cell accumulation and accelerates atherosclerosis in mice. J. Clin. Invest. 117, 3900–3908 (2007).

66. Sturek, J. M. et al. An intracellular role for ABCG1-mediated cholesterol transport in the regulated secretory pathway of mouse pancreatic beta cells. J. Clin. Invest. 120, 2575– 2589 (2010).

67. Shimano, H. & Sato, R. SREBP-regulated lipid metabolism: convergent physiology - divergent pathophysiology. Nat. Rev. Endocrinol. 13, 710–730 (2017).

68. Diraison, F. et al. SREBP1 is required for the induction by glucose of pancreatic beta-cell genes involved in glucose sensing. J. Lipid Res. 49, 814–822 (2008).

69. Lee, G. et al. SREBP1c-PAX4 Axis Mediates Pancreatic β-Cell Compensatory Responses Upon Metabolic Stress. Diabetes 68, 81–94 (2019).

70. Cai, X. et al. Association between prediabetes and risk of all cause mortality and cardiovascular disease: updated meta-analysis. BMJ 370, m2297 (2020).

71. Yuan, J.-M., Stram, D. O., Arakawa, K., Lee, H.-P. & Yu, M. C. Dietary cryptoxanthin and reduced risk of lung cancer: the Singapore Chinese Health Study. Cancer Epidemiol. Biomark. Prev. Publ. Am. Assoc. Cancer Res. Cosponsored Am. Soc. Prev. Oncol. 12, 890–898 (2003).

72. Wang, Y.-L., Koh, W.-P., Yuan, J.-M. & Pan, A. Plasma ferritin, C-reactive protein, and risk of incident type 2 diabetes in Singapore Chinese men and women. Diabetes Res. Clin. Pract. 128, 109–118 (2017).

73. Chan, S. H. et al. Analysis of clinically relevant variants from ancestrally diverse Asian genomes. Nat. Commun. 13, 6694 (2022).

74. Aryee, M. J., et al. Minfi: a flexible and comprehensive Bioconductor package for the analysis of Infinium DNA methylation microarrays. Bioinforma. Oxf. Engl. 30, 1363– 1369 (2014).

75. Lehne, B. et al. A coherent approach for analysis of the Illumina HumanMethylation450 BeadChip improves data quality and performance in epigenome-wide association studies. Genome Biol. 16, 37 (2015).

76. Houseman, E. A. et al. DNA methylation arrays as surrogate measures of cell mixture distribution. BMC Bioinformatics 13, 86 (2012).

77. Willer, C. J., Li, Y. & Abecasis, G. R. METAL: fast and efficient meta-analysis of genomewide association scans. Bioinforma. Oxf. Engl. 26, 2190–2191 (2010).

78. Kujala, U. M. et al. Polygenic Risk Scores and Physical Activity. Med. Sci. Sports Exerc. 52, 1518–1524 (2020).

79. Privé, F. et al. Portability of 245 polygenic scores when derived from the UK Biobank and applied to 9 ancestry groups from the same cohort. Am. J. Hum. Genet. 109, 12– 23 (2022).

80. Tanigawa, Y. et al. Significant sparse polygenic risk scores across 813 traits in UK Biobank. PLoS Genet. 18, e1010105 (2022).

81. Ge, T. et al. Development and validation of a trans-ancestry polygenic risk score for type 2 diabetes in diverse populations. Genome Med. 14, 70 (2022).

82. Lam, M. et al. Collective genomic segments with differential pleiotropic patterns between cognitive dimensions and psychopathology. Nat. Commun. 13, 6868 (2022).

83. Purcell, S. et al. PLINK: a tool set for whole-genome association and population-based linkage analyses. Am. J. Hum. Genet. 81, 559–575 (2007).

84. Roadmap Epigenomics Consortium et al. Integrative analysis of 111 reference human epigenomes. Nature 518, 317–330 (2015).

85. Hammal, F., de Langen, P., Bergon, A., Lopez, F. & Ballester, B. ReMap 2022: a database of Human, Mouse, Drosophila and Arabidopsis regulatory regions from an integrative analysis of DNA-binding sequencing experiments. Nucleic Acids Res. 50, D316–D325 (2022).

86. Shabalin, A. A. Matrix eQTL: ultra fast eQTL analysis via large matrix operations. Bioinforma. Oxf. Engl. 28, 1353–1358 (2012).

87. Kolberg, L., Raudvere, U., Kuzmin, I., Vilo, J. & Peterson, H. gprofiler2 -- an R package for gene list functional enrichment analysis and namespace conversion toolset g:Profiler. F1000Research **9**, ELIXIR-709 (2020).

88. Yew, Y. W. et al. Investigating causal relationships between obesity and skin barrier function in a multi-ethnic Asian general population cohort. Int. J. Obes. 2005 47, 963– 969 (2023).

89. Kwong, A. M. et al. Robust, flexible, and scalable tests for Hardy-Weinberg equilibrium across diverse ancestries. Genetics 218, iyab044 (2021).

90. Yang, J., Lee, S. H., Goddard, M. E. & Visscher, P. M. GCTA: a tool for genome-wide complex trait analysis. Am. J. Hum. Genet. 88, 76–82 (2011).

91. Yang, J. et al. Common SNPs explain a large proportion of the heritability for human height. Nat. Genet. 42, 565–569 (2010).

92. Sadhu, N. et al. Metabolome-wide association of carotid intima media thickness identifies FDX1 as a determinant of cholesterol metabolism and cardiovascular risk in Asian populations. Preprint at 10.1101/2024.05.14.24307316 (2024).

93. Mbatchou, J. et al. Computationally efficient whole-genome regression for quantitative and binary traits. Nat. Genet. 53, 1097–1103 (2021).

94. Castro-Mondragon, J. A. et al. JASPAR 2022: the 9th release of the open-access database of transcription factor binding profiles. Nucleic Acids Res. 50, D165–D173 (2022).

95. Grant, C. E., Bailey, T. L. & Noble, W. S. FIMO: scanning for occurrences of a given motif. Bioinforma. Oxf. Engl. 27, 1017–1018 (2011).

96. Bailey, T. L., Johnson, J., Grant, C. E. & Noble, W. S. The MEME Suite. Nucleic Acids Res. 43, W39–49 (2015).

97. Amemiya, H. M., Kundaje, A. & Boyle, A. P. The ENCODE Blacklist: Identification of Problematic Regions of the Genome. Sci. Rep. 9, 9354 (2019).

98. Quinlan, A. R. & Hall, I. M. BEDTools: a flexible suite of utilities for comparing genomic features. Bioinforma. Oxf. Engl. 26, 841–842 (2010).

99. Picard Tools - By Broad Institute. https://broadinstitute.github.io/picard/.

100. McAllan, L. et al. Integrative genomic analyses in adipocytes implicate DNA methylation in human obesity and diabetes. Nat. Commun. 14, 2784 (2023).

101. Tibshirani, R. Regression Shrinkage and Selection via the Lasso. J. R. Stat. Soc. Ser. B Methodol. 58, 267–288 (1996).

