## Supplementary Figures for "Nuclear regulatory disturbances precede and predict the development of Type-2 diabetes in Asian populations"

### Table of Contents

#### Supplementary Figures


| SF | NAME | PAGE |
|---|---|---|
| **1** | Sensitivity analyses | 2 |
| **2** | Gene set enrichment analysis (*cis*-eQTMs) | 3 |
| **3** | Gene set enrichment analysis (*trans*-eQTMs) | 4 |
| **4** | Hierarchical Clustering of *trans*-mQTLs | 5 |
| **5** | Sankey plots for *trans*-mQTL clusters | 6 – 7 |
| **6** | *PANK1* and sentinel CpGs | 8 |
| **7** | *SCARB1* and sentinel CpGs | 9 |

**Supplementary Figure 1: Comparisons of association models without and with adjustment for potential confounding variables**. Scatter plot showing the effect size estimates in the base model (CpG + Covariates), compared to the model adjusted for potential confounding variables including **(a**) glucose, **(b)** Hba1C, **(c)** Glucose + Hba1C, **(d)** BMI + **(e)** top *cis*-mQTL, and **(f)** base model with people with prediabetes excluded. The sample size (N), pearson's correlation coefficient (r) and the p-value of correlation (P) are labelled.

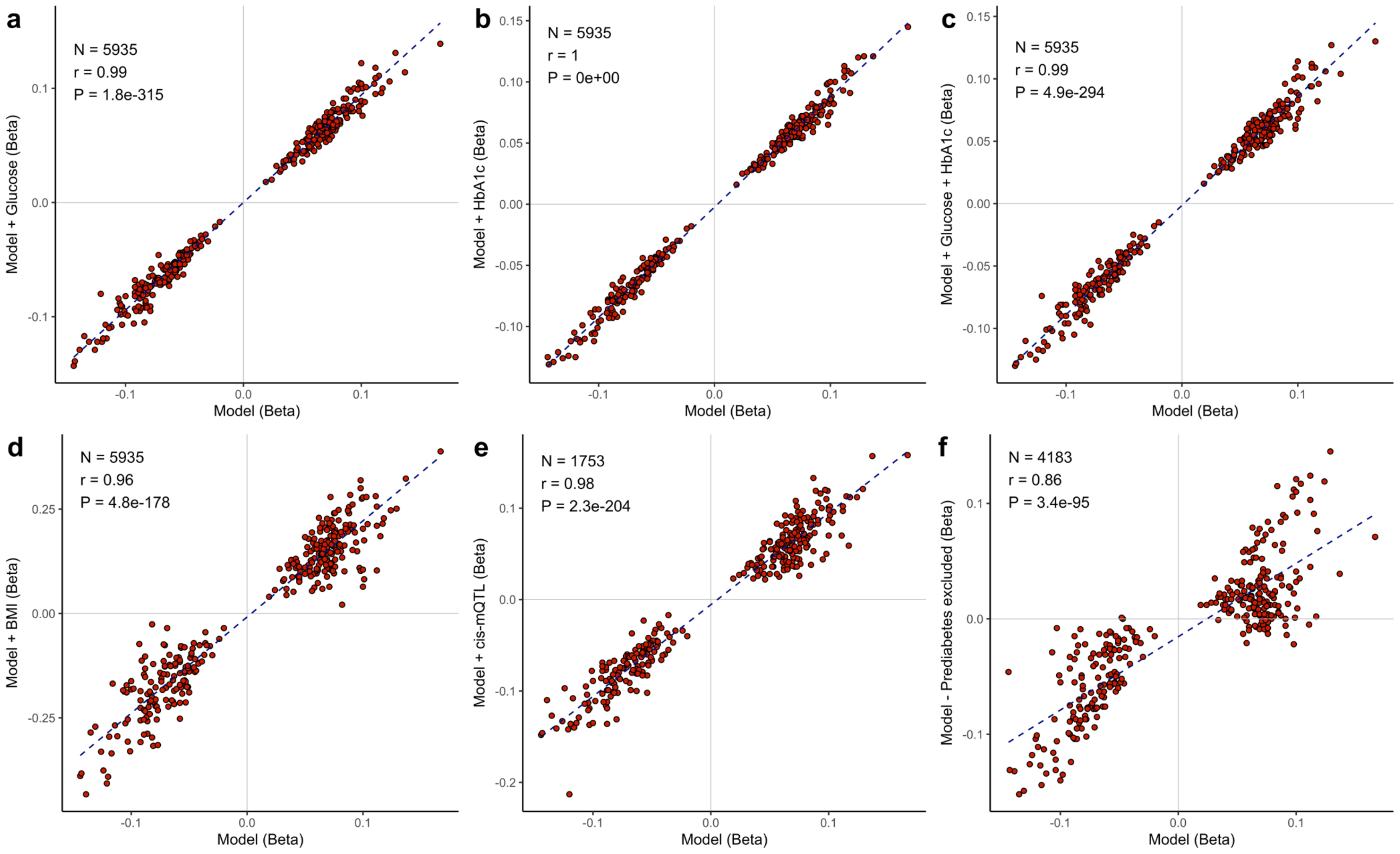

**Supplementary Figure 2: Gene set Enrichment analysis of *cis*-eQTMs.** The plot shows the enrichment analyses of the 24 *cis*-eQTM genes significantly associated with the sentinel CpGs. Enrichment was performed for the different terms in the four pathway databases (Gene Ontology, KEGG, Reactome and Wiki pathways) using the g: profiler tool. The x-axis is the term name, and the y-axis represents the -log10 (P) value for enrichment estimated using hypergeometric testing for overrepresentation. The colors indicate non-significant (grey), nominally significant (light blue), and significant associations after multiple testing correction (navy).

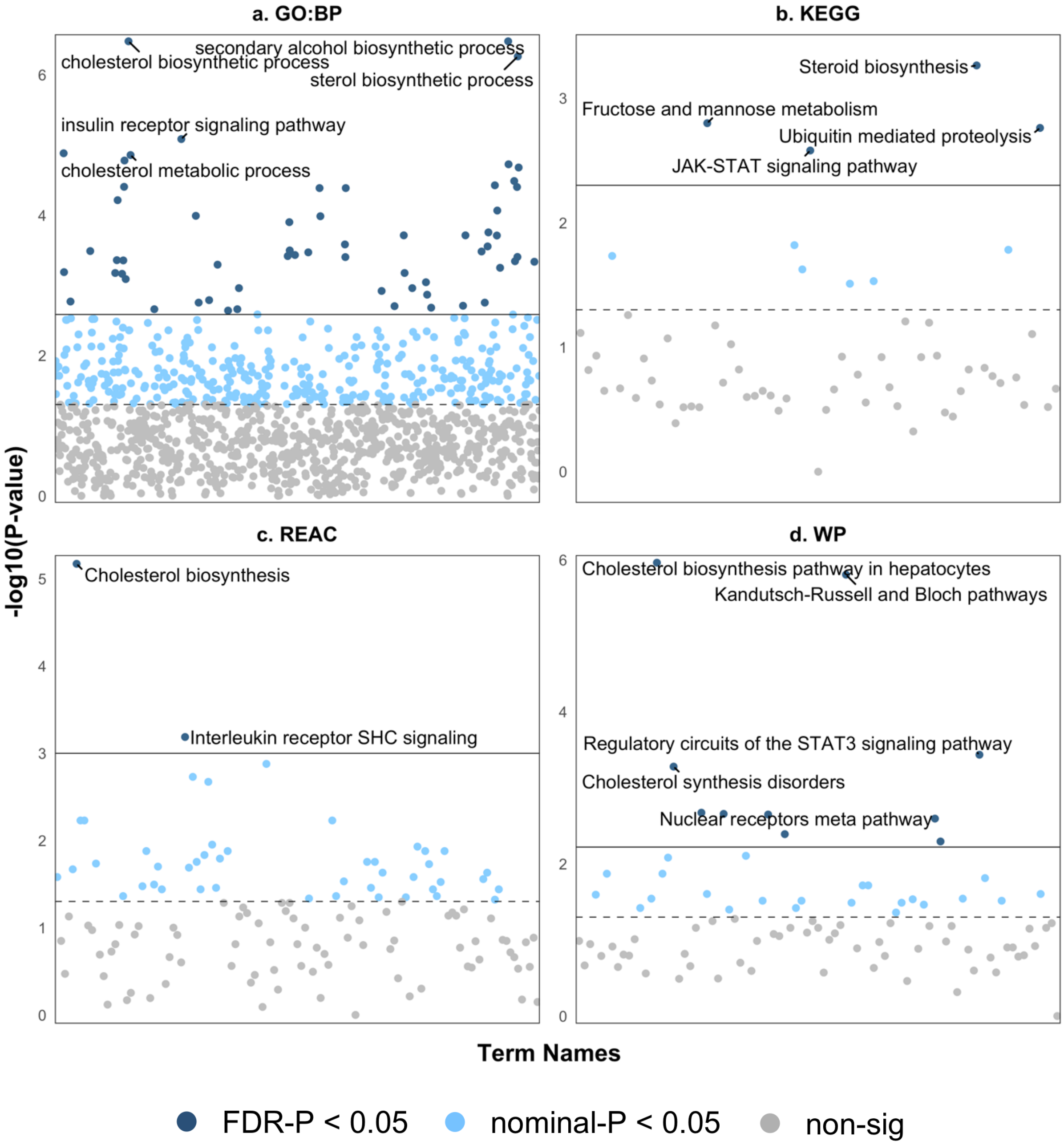

**Supplementary Figure 3: Gene set Enrichment analysis of *trans*-eQTMs.** The plot shows the enrichment analyses of the 764 *trans*-eQTM genes significantly associated with the sentinel CpGs. Enrichment was performed for the different terms in the four pathway databases (Gene Ontology, KEGG, Reactome and Wiki pathways) using the g: profiler tool. The x-axis is the term name, and the y-axis represents the -log10 (P) for enrichment estimated using hypergeometric testing for overrepresentation. The colors indicate non-significant (grey), nominally significant (light blue), and significant associations after multiple testing correction (navy).

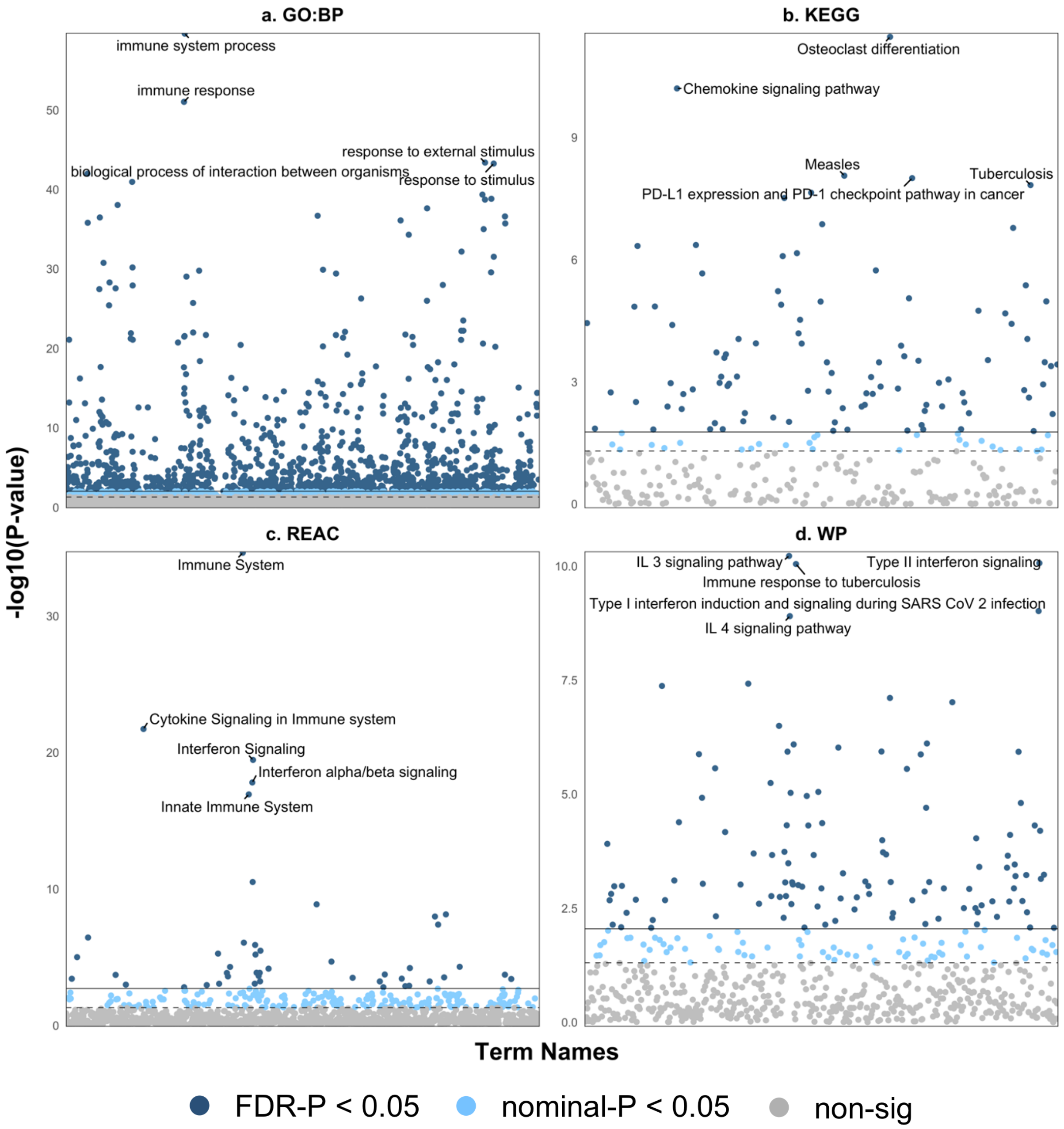

**Supplementary Figure 4: Hierarchical clustering of the trans-mQTLs.** The 33 SNPs with at least one significantly associated cis-eQTLs were clustered into seven sets based on their shared CpGs. The clustering was performed using the dissimilarity matrix applying the average linkage method. (a) Average Silhouette width scores for cluster sizes from two to 15. This was used to select the optimal number of clusters. (b) the dendrogram showing the organization of the 33 SNPs and the boxes highlight the defined clusters at K=7.

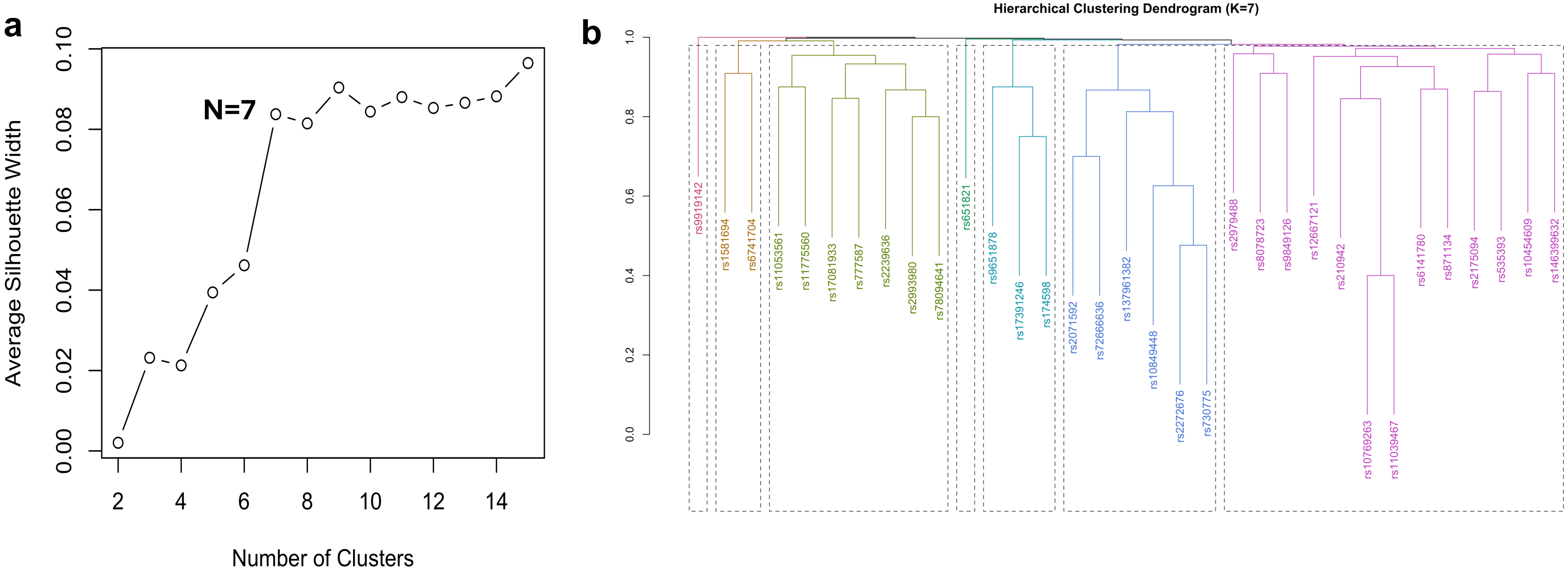

**Supplementary Figure 5: Sankey Plots for the *trans*-mQTL clusters.** The Plot shows the seven identified *trans*-acting mQTL clusters and their associated genes and CpGs. The first layer are the SNPs; the second layer are the significantly associated eQTL genes for each SNP and the final layer are significantly associated and colocalized trans-CpGs and their nearest genes.

**Cluster 1: 13 mQTLs and 83 CpGs**

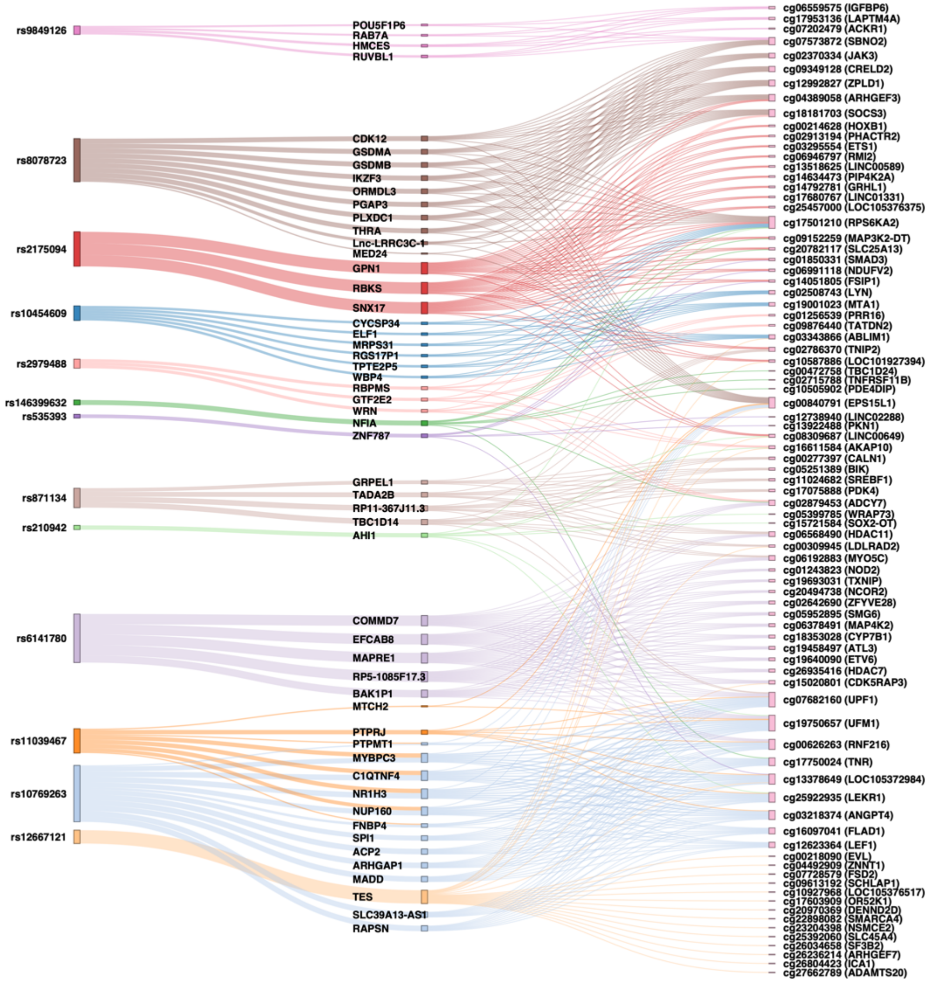


**Cluster 2: 6 mQTLs and 58 CpGs**

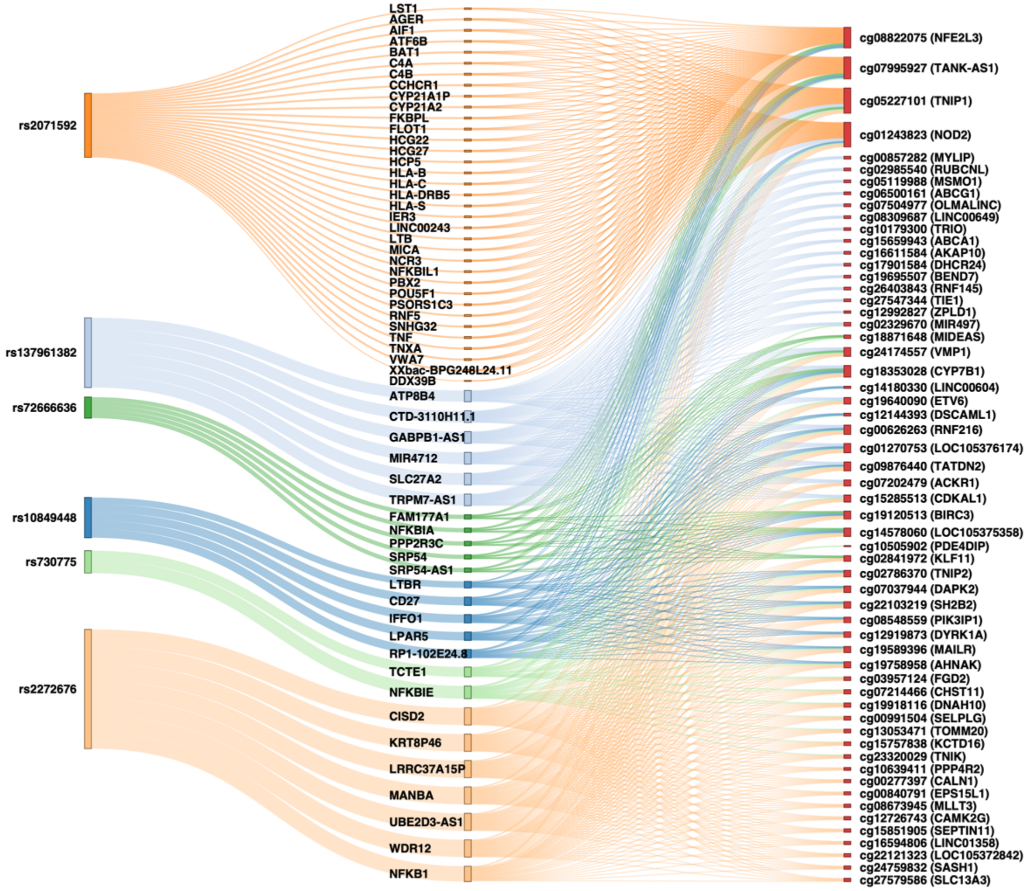

### Cluster 3: 7 mQTLs and 42 CpGs

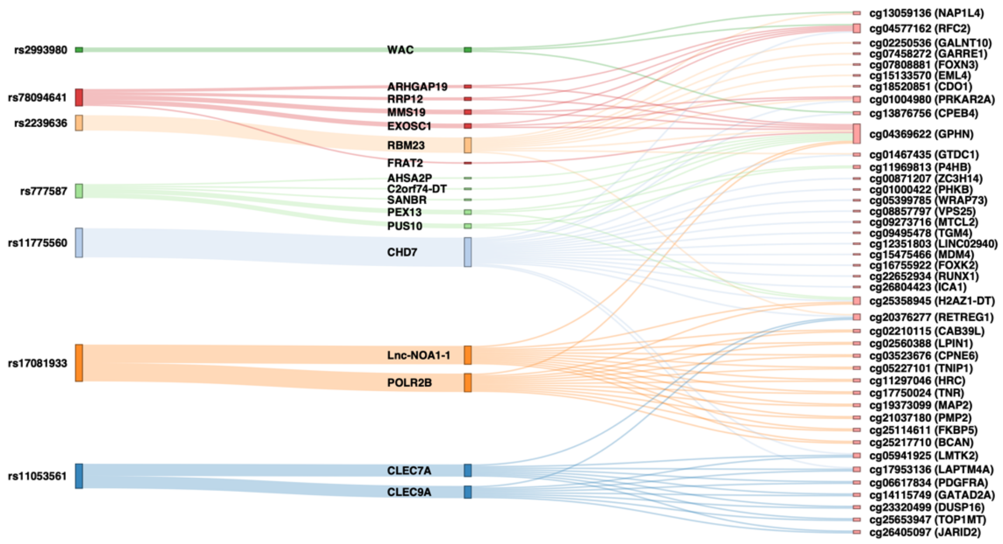


### Cluster 4: 2 mQTLs and 11 CpGs

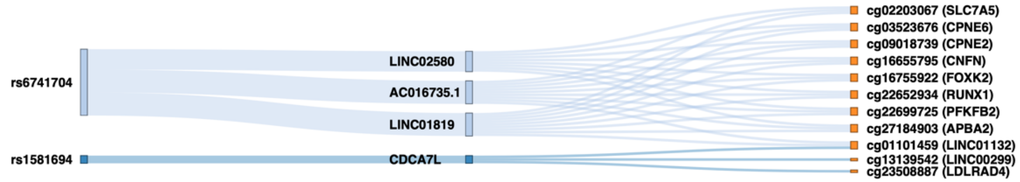


### Cluster 5: 3 mQTLs and 5 CpGs

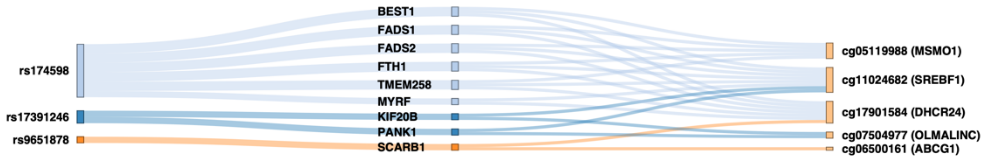


### Cluster 6: 1 mQTLs and 2 CpGs

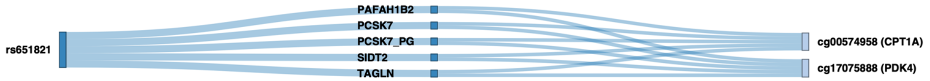


### Cluster 7: 1 mQTLs and 2 CpGs

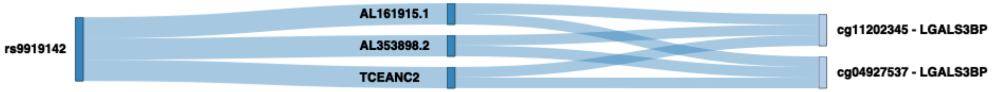

**Supplementary Figure 6: *PANK1* and *trans*-activation of Sentinel CpGs.** Regional association plot showing the association of the region around the lead SNP rs17391246(A>G) with two T2D associated sentinel CpGs in *trans*, cg05704997 (*OLMALINC*) and cg11024682 (*SREBF1*). SMR and colocalization was performed between *PANK1* expression (highlighted cis-eQTL) and DNA methylation at the two CpG loci. The SMR value is the effect size and Standard error estimated of association.

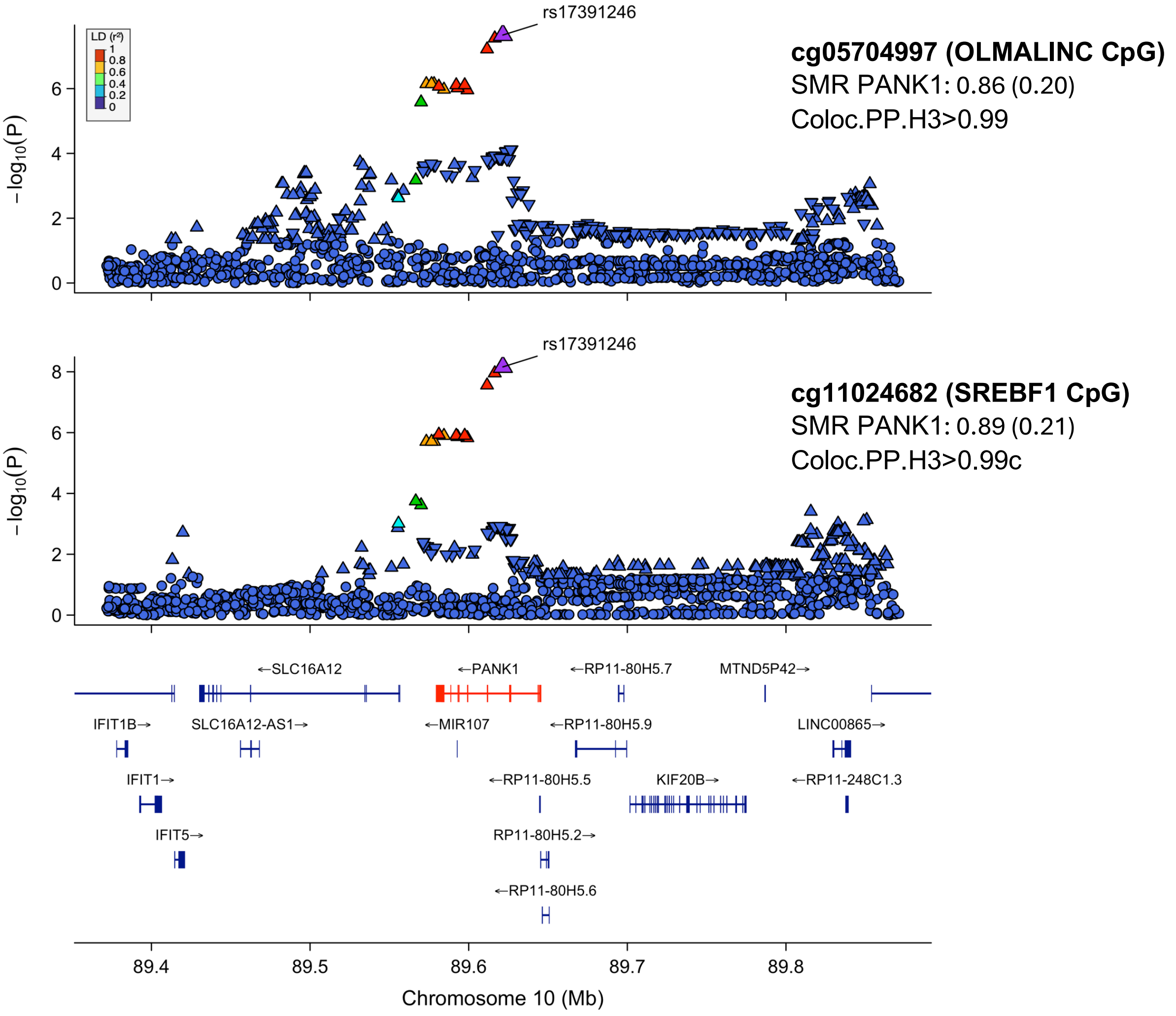

**Supplementary Figure 7: *SCARB1*, Sentinel CpGs, Total Cholesterol and HDL Cholesterol.** Regional association plot showing the association of the region around the lead SNP rs9651878(A>G) with two T2D associated sentinel CpGs cg06500161 (*ABCG1*), cg17901584 (*DHCR24*), total cholesterol, and HDL cholesterol. SMR and colocalization was performed between *SCARB1* expression (highlighted cis-eQTL) and DNA methylation at the two CpG loci, HDL cholesterol and total cholesterol levels. The SMR value is the effect size and Standard error estimated of association. The Sentinel *trans*-acting SNP (rs9651878) is labelled in all plots. In addition, the strongest associated SNP for TC and HDL at the *SCARB1* loci are identified (rs921919 and rs10773112 respectively). SNP rs9651878 is in low LD with both rs921919 and rs10773112 ($R^2$<0.001), suggesting its effect on TC and HDL is independent from these variants.

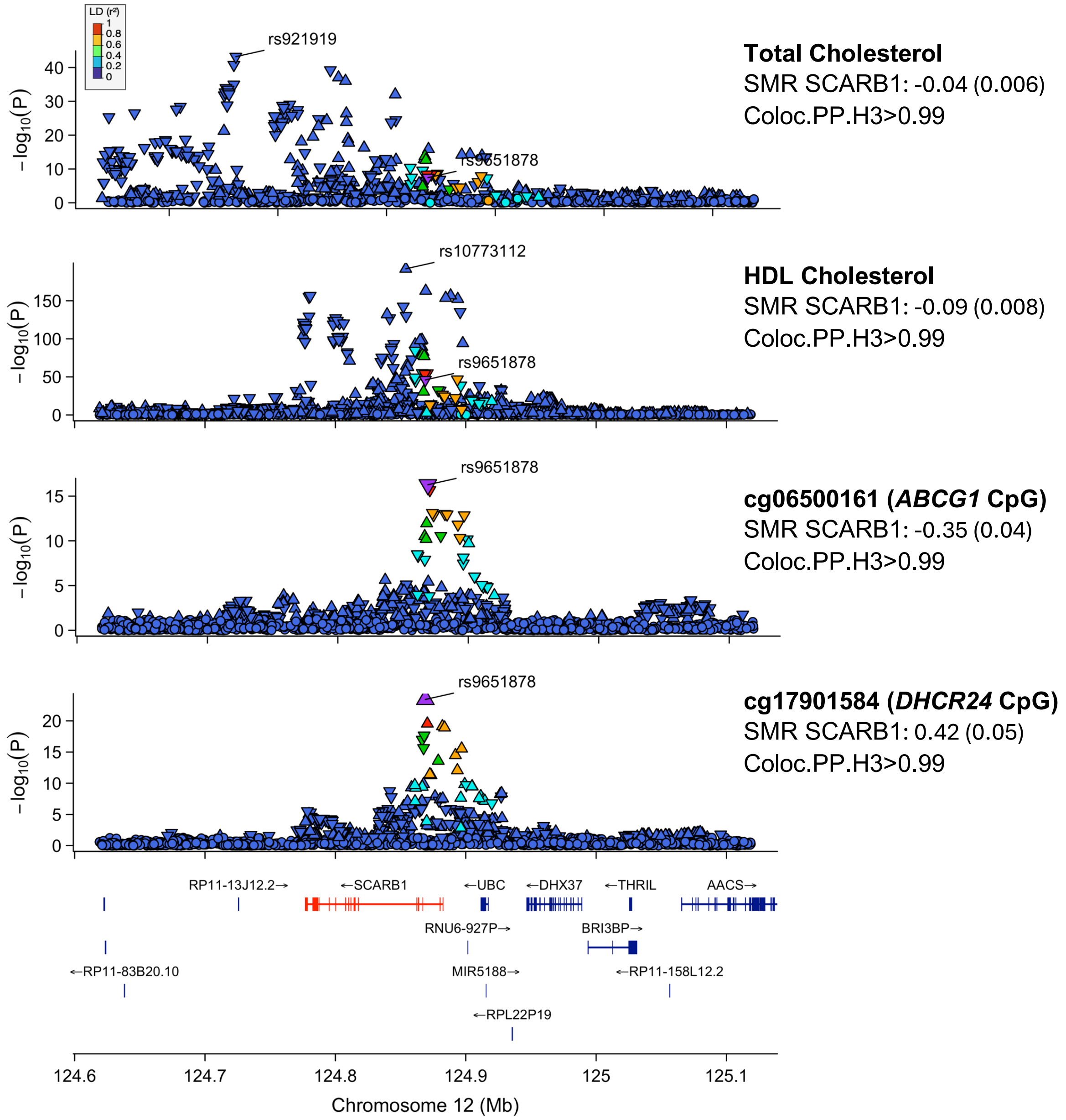
